# Pretrained transformers applied to population cancer registries improve survival prediction in label-scarce and previously unseen cancers

**DOI:** 10.64898/2026.08.30.26361693

**Authors:** Yuzhen Gao, Shaobo Yu, Yan Xia, Shipeng Chen, Shenglong Xia, Rui An, Jing Zeng, Feng Zhao, Yilei Ma, Yanzhong Wang, Xinyou Xie, Jun Zhang

## Abstract

Prognostic models in oncology are developed one cancer at a time, from that cancer’s own labelled outcomes, and fail where prognostic information is scarcest. Rare cancers account for roughly a fifth of diagnoses and most paediatric malignancies, yet seldom supply enough events for a reliable time-to-event model. We therefore asked whether a representation learned without outcome labels can supply what those cohorts cannot. A Transformer encoder was pretrained by masked field-value modelling on 9,425,135 tumour records from the SEER 17 registries, diagnosed in 2000–2023. Only diagnosis-time fields passing a fail-closed coding-verification gate were admitted, and each record was emitted as an era-specific and a harmonised view, keeping two decades of recoding auditable. The encoder was then frozen and read by a linear Cox head for overall survival. Nine rare cancers were removed from the pretraining corpus entirely, each requiring an independent pretraining run. On a sealed test partition, all nine exceeded an architecture-identical random frozen encoder in Harrell concordance by +0.0034 to +0.0368, every lower confidence limit above zero. At 256 labelled patients, all 67 cancers favoured the pretrained representation over budget-matched Cox regression, median difference +0.0283. The advantage was bounded: given the entire training set, Cox regression was favoured in seven of nine rare cancers. The encoder did not outperform a field-frequency baseline on its own objective, so upstream reconstruction did not predict downstream transfer. Outcome-agnostic registry pretraining carries prognostic signal into cancers it has never seen, and is most useful where labels are fewest, without establishing clinical utility.

## Introduction

Prognosis after a cancer diagnosis is estimated, in routine practice, from a small and highly structured set of registry-coded facts: primary site, histology, grade, stage, tumour size and age. Population-based cancer registries record precisely these variables for entire populations and follow patients for vital status, which makes them the principal substrate for population-level survival evidence and for the prognostic models built upon it [1]. The problem they cover is not small: close to 20 million new cancer cases and 9.7 million cancer deaths were estimated worldwide for 2022 [2]. The modelling convention applied to these data, however, has remained resolutely local. One model is fitted per cancer, from the labelled outcomes of that cancer alone, and nothing learned in one disease is carried into the next.

That convention degrades exactly where prognostic information is scarcest. Rare cancers, conventionally defined by an incidence below six cases per 100,000 persons per year, account for approximately 20% of cancer diagnoses in the United States, comprise 71% of cancers in children and adolescents, and carry substantially poorer five-year relative survival than common cancers, at 55% versus 75% in males and 60% versus 74% in females [3,4]. Their individual cohorts nevertheless contain few patients and fewer events, and commonly fall below the sample size at which a multivariable time-to-event model can be developed with acceptable optimism and precision [5]. The cancers with the greatest need for a reliable prognostic model are therefore the cancers least able to supply the labels needed to fit one, and that shortfall is clinical rather than merely methodological: the evidence is thinnest precisely where the survival deficit is largest and the affected population youngest [3,4].

Self-supervised pretraining is the obvious candidate for closing that gap. Transformer encoders trained by masked reconstruction learn representations from unlabelled data that transfer to downstream tasks with few labels [6,7], and the same paradigm has been proposed as the organising principle for generalist medical models [8]. In structured health data, encoders pretrained on sequences of diagnosis codes or phenotypic traits have yielded representations that transfer across clinical endpoints [9,10], including models that tokenise thousands of individual-level biobank traits without positional encoding [11] and frameworks that predict many diseases from one shared representation [12]. Self-supervised pretraining on longitudinal event streams has been extended to the time-to-event setting itself, improving label efficiency across many outcomes and across institutions [13], and a transformer over clinicogenomic feature-value pairs, pretrained by self-supervision and adapted by transfer learning, improves survival prediction in trial cohorts [14]. In general tabular prediction, a single pretrained network can outperform tuned task-specific pipelines on small datasets [15]. The shared premise is that the statistical dependencies linking clinical fields are partly disease-agnostic, so a representation learned once across many patients may substitute for labels that a single small cohort cannot provide.

Whether that premise holds for cancer registry data is unresolved, for three reasons. First, evidence for structured-data foundation models rests largely on comparisons against task-specific baselines on random splits of a single population, and rarely against an architecture-identical randomly initialised encoder; the contribution of pretraining therefore cannot be separated from the contribution of the architecture or of the input encoding, and reported gains have been argued to outrun the evaluations that support them [16]. Second, registry variables are not stationary. Coding schemes are revised, and the 2018 adoption of the eighth edition of the AJCC staging system, together with the corresponding registry recodes for stage, grade and tumour size, means that a field carrying the same name in 2003 and in 2020 need not carry the same meaning [17]. A model that collapses a multi-decade registry into one encoding mixes those definitions silently and cannot be audited afterwards. Third, cross-cancer transfer has been examined, but under assumptions that differ from the registry setting. In molecular cohorts, pan-cancer pretraining with fine-tuning on a held-out tumour type, meta-learning across tasks, and a mixture-of-experts framework have each improved survival prediction in small or unseen tumour types [18,19,20]; in registry data, supervised multitask learning across ten HPV-associated cancers improved five-year survival prediction by sharing information between anatomically distinct sites [21]. Each pretrains or trains with outcome labels, and all but the last use molecular rather than registry inputs. What remains less established is the transfer question that matters most in the registry setting: whether a representation learned without any outcome label, from other cancers only, assists a cancer that contributed nothing to pretraining. Holding an entire disease out of pretraining requires retraining the encoder once per disease, which is why the question is rarely posed in this form.

We addressed these three problems in a single pan-cancer registry cohort. SEERFound is a Transformer encoder pretrained by masked field-value modelling on 9,425,135 eligible tumour records diagnosed between 2000 and 2023 in the SEER 17 registries, using only diagnosis-time fields that passed a fail-closed categorical-coding verification gate. Cross-era drift is handled explicitly rather than by reconciliation: each record is emitted as two aligned views, an era-specific raw view and a harmonised canonical view built from official registry recodes, so that era-dependent and era-invariant encodings remain separately visible to the model. The encoder is then frozen, and a linear Cox head is fitted on top of it for overall survival within each cancer separately. Three independent questions were specified in advance: how the encoder performs on diagnostics of its masked-field pretraining objective relative to a field-frequency baseline; whether it aligns the two views of the same tumour and yields more reproducible cluster structure than an architecture-identical random encoder; and whether it improves overall-survival prediction against a random frozen encoder, a budget-matched Cox model and the same architecture trained end-to-end, including in nine rare cancers each removed entirely from the pretraining corpus and re-pretrained from scratch. The setting this design targets is the cancer for which no disease-specific model can be developed at all, so the analyses are framed to show not only whether pretraining helps but at what label budget its advantage ends. We report the direction and uncertainty of each analysis without making the interpretation of one conditional on another.

## **1.** Methods

### **1.1** Study design and objectives

This retrospective population-registry study first pretrained a Transformer encoder on diagnosis-time fields without outcome labels, then evaluated the frozen representation for overall survival (OS) within individual cancers. Each research question had a written protocol specifying its comparator set and metric interpretation before the corresponding results were generated; Supplementary Methods S2 documents the protocols and their information state at freezing. Downstream analyses were indexed by the number of labelled patients available, making the point at which pretraining ceased to be preferable directly observable. Reporting follows STROBE, RECORD and TRIPOD+AI [22–24], with completed item-by-item checklists in the Supplementary Reporting Checklists.

### **1.2** Data source and control of the source snapshot

The analysis used one frozen extract of the SEER Research Plus 17-registry file, November 2025 submission, covering diagnosis years 2000–2023 and containing 9,708,868 tumour records and 250 fields [25]. The matching variable dictionary, Data Description and official SEER recode pages governed field semantics. All reported counts and checks used the complete extract; no simulated, imputed or placeholder data were used. Supplementary Methods S1 lists the documentary sources and checksums.

### **1.3** Eligibility

Eligible records had behaviour coded as Malignant under both ICD-O-2 and ICD-O-3, a reporting source other than death certificate only or autopsy only, and valid age, primary site, histology and diagnosis year. The era-limited malignant categories were not merged into the cohort definition. Criteria were evaluated marginally; exclusion and cohort counts are reported in Section 2.1 and Figure 1A.

**Figure 1.**
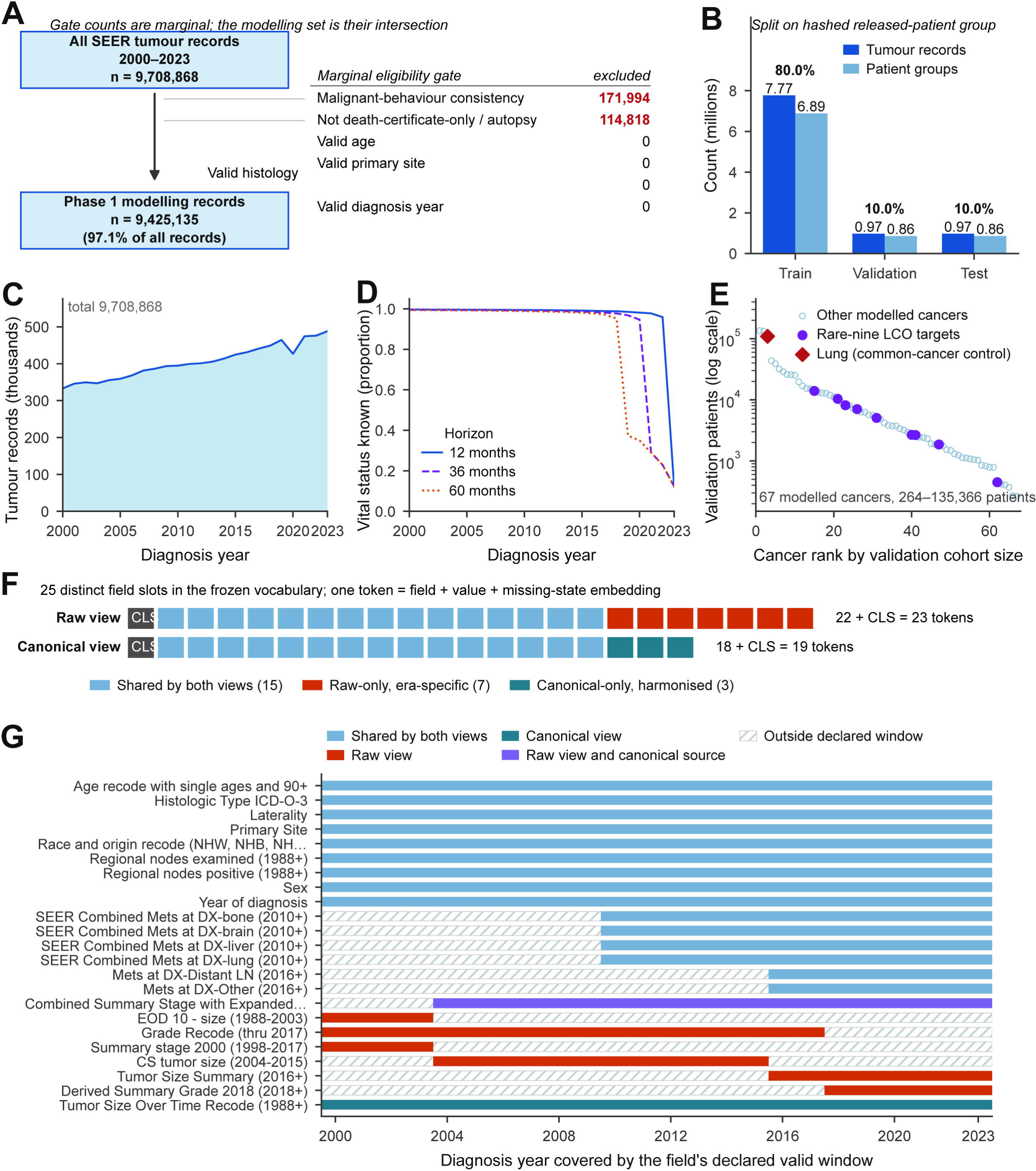
Study cohort, temporal structure, and dual-view input. A, Cohort assembly from 9,708,868 SEER tumour records diagnosed in 2000–2023; exclusion counts are marginal and the modelling cohort is their intersection. B, Patient-group hash split into training, validation, and test sets (80:10:10), preventing patient crossover. C, Annual tumour-record counts. D, Proportion with known vital status at 12, 36, and 60 months by diagnosis year. E, Validation-cohort sizes for 67 cancers; purple circles mark the nine leave-cancer-out (LCO) targets and the red diamond marks lung cancer as a common-cancer control. F, Raw and canonical token layouts; colours distinguish shared, raw-only, and canonical-only fields. G, Declared diagnosis-year windows for selected fields; hatching denotes years outside a field’s valid window. CLS, classification token; LCO, leave-cancer-out.

### **1.4** Verification of categorical coding

Before any categorical field, event, reference group or cancer target was defined, a fail-closed gate verified the release-specific value labels, observed ranges, missing codes and diagnosis-year frequencies, then tested each cited raw-to-canonical mapping by cross-tabulation and impossible-mapping checks. A frozen specification recorded the role, view, valid-year window, missing semantics, leakage rule and source of all 250 fields; its concept map covered 3,620 observed values. Of these fields, 23 diagnosis-time inputs and 2 filter-only fields were admitted. Fine-grained AJCC/EOD T, N and M fields, site-specific factors and biomarkers were excluded because era-consistent coding could not be verified. Sixteen checks for coverage, conservation, leakage and patient-identifier use passed. Supplementary Methods S3 gives the checks and resolved coding decisions.

### **1.5** Cross-era harmonisation and the dual-view input

Because 13 of 23 inputs are not defined throughout 2000–2023, each record was represented by aligned raw and canonical token sequences. The raw view retains the era-specific grade, stage and size fields populated in the record’s diagnosis year. The canonical view substitutes a verified partial four-level grade mapping, broad summary stage and SEER Tumour Size Over Time recode. Official recodes were used where available; no site-specific legal-range or midpoint rule was derived locally. Stage and size crosswalks covered all 9,708,868 source records, with zero broad-stage contradictions among 2004–2017 records in which both stage fields were present. Supplementary Table S2 gives each slot and valid-year window.

The raw and canonical views contain 22 and 18 field tokens, respectively, plus CLS. Tokens sum field-identity, value and missing-state embeddings; structural era-blanks remain distinct from Unknown and Not applicable. No positional encoding was used, non-CLS tokens were shuffled, and the vocabulary was fitted only on the training split. Complete vocabulary and token-construction details are in Supplementary Methods S4.

### **1.6** Outcome definition and follow-up maturity

OS was measured from the index diagnosis using SEER Survival months. Unknown time was neither converted nor imputed; valid time required an accepted complete- or incomplete-date survival flag. Death was defined by Vital status recode (study cutoff used), under which deaths after the study cut-off are recoded as alive at cut-off; alive records were censored. Records flagged Not calculated were death-certificate-only or autopsy cases and were excluded. Outcome, treatment, identifier and post-diagnosis fields were blocked from the vocabulary and rechecked at every training preflight.

Follow-up maturity was evaluated by diagnosis year. Fixed-horizon metrics were restricted to years with empirical support: diagnosis through 2022 for 12 months, through 2020 for 36 months and through 2018 for 60 months; 2023 contributed only to self-supervised training. Full outcome-validation counts and maturity rules are in Supplementary Methods S3 and S10.

### **1.7** Data partitioning

Whole patient groups were assigned to training, validation and test in an 80/10/10 ratio by a deterministic salted hash of the released patient identifier. Cross-split crossover was zero; identifiers were used only for grouping and were not persisted. Emitting two views from 9,425,135 eligible records produced 18,850,270 sequences, including 15,079,716 from 7,539,858 training records. Split-level record, patient-group and sequence counts are reported in Section 2.1 and Supplementary Figure S1. The case-level export lacks a registry identifier, precluding registry holdout and domain-adversarial designs.

### **1.8** Self-supervised pretraining

Pretraining used masked field-value modelling [7], alternating random masking of 15% of non-CLS fields with semantic-block masking of stage, metastasis, grade, size or morphology. Selected positions were replaced by a mask token with probability 0.8, a random in-field value with probability 0.1 and left unchanged otherwise. Per-field softmax heads predicted only values legal for the masked field. The encoder was an 8-layer, 8-head pre-norm Transformer [6] with width 256 and no positional encoding. AdamW optimisation used a learning rate of 3 × 10□□, weight decay 0.01, micro-batches of 1,024 with four accumulation steps, three epochs and seed 20260802. The run ended at optimizer step 11,084; that terminal checkpoint was used unchanged in every downstream analysis, without checkpoint selection or further tuning. Supplementary Methods S5 gives the complete configuration, telemetry and pilot checks.

### **1.9** Diagnostic evaluation of the pretraining objective

The frozen encoder was compared with the training-set field-frequency baseline under deterministic paired raw/canonical masking on the held-out test partition. Negative log-likelihood (NLL; lower is better) was the primary diagnostic metric; macro F1 and rare-class Recall@5 were secondary and descriptive. The reference comparison required the upper 95% confidence limit of model-minus-baseline NLL to fall below zero.

The reporting order was frozen before any reconstruction prediction existed, but after pretraining and after other transfer results had been read; the plan is therefore late prospective, not preregistered or fixed before model development. None of the study protocols was deposited with an independent timestamp, so prespecified and protocol-frozen mean only that the written protocol existed before the result it governed. The reconstruction diagnostic neither conditions nor acts as a gate for downstream analyses. The subsequent 40-target field decomposition was descriptive. Protocol chronology and claim limits are detailed in Supplementary Methods S2 and S13.

### **1.10** Evaluation of representation structure and stability

Both analyses used validation data only. Bidirectional cross-view retrieval compared L2-normalised CLS embeddings from 10,000 patient groups by Recall@1, Recall@5, mean reciprocal rank (MRR) and mean rank, with 10,000 paired bootstrap resamples. Its reference point required the lower 95% limit versus the random encoder to exceed zero for both Recall@1 and MRR.

Clustering stability applied MiniBatchKMeans at K = 8, 16 and 32 to 20,000 patient groups, with 30 paired 80%-subsample repeats per K and adjusted Rand index (ARI) agreement. Its reference point required lower limits above zero for at least two K values. The two branches and reference points were fixed and reported together.

### **1.11** Downstream survival analysis

#### 1.11.1 Cancer targets and index-tumour definition

Targets used the ICD-O-3/WHO 2008 site recode, except paediatric neuroblastoma, defined by ICCC-3/IARC 2017 category IVa at ages 0–19. A result-independent rule included site values with at least 1,024 eligible training patients, the largest protocol-frozen budget. Values equal to Miscellaneous, beginning with Other, or ending in NOS were removed as administrative categories before expansion results were read. Hodgkin–extranodal (919 training patients) and pleura (452) fell below the threshold. The final panel comprised 67 targets; Supplementary Table S1 gives verified definitions and counts.

For each target, the index tumour was the earliest eligible diagnosis, with ties resolved by source-row order. One row per patient group and target was enforced. A patient could enter more than one target cohort but never more than one split.

#### 1.11.2 Primary representation contrast

The primary contrast was the pretrained frozen encoder minus an architecture-identical random frozen encoder. Both used a fixed non-affine output LayerNorm, received the canonical view and the same labelled patients, order, seed, horizons and evaluation rows, and trained only a zero-initialised 256-parameter linear Cox head. Encoder weights were frozen and placed in evaluation mode in both arms. The complete-risk-set Cox partial likelihood used Efron tie handling [26,27], never a mini-batch risk set, and was checked at run time against an independent direct calculation. This contrast establishes whether a linear head can read signal created by pretraining; by itself it establishes neither superiority to conventional or end-to-end models nor transfer to unseen cancers. Probe settings and checks are in Supplementary Methods S6.

#### 1.11.3 Label-budget protocol

Budgets were 32, 64, 128, 256, 512 and 1,024 unique patient groups, with 20 nested, deterministic repeats per target and identical patients across arms. Each of 1,000 paired hierarchical-bootstrap replicates sampled a repeat and then its evaluation patient groups, propagating labelled-subset and evaluation-sample variability. Formal development and 67-target test classifications used the mean observed repeat difference; the validation budget panel and rare-nine test extension plotted the bootstrap median. Supplementary Methods S8 and S11 specify sampling, rejected partial grids and interval construction.

#### 1.11.4 Leave-cancer-out pretraining

Nine rare cancers were held out in nine independent pretraining runs: acute myeloid leukaemia, chronic lymphocytic leukaemia, myeloma, nodal Hodgkin lymphoma, mesothelioma, salivary gland, soft tissue including heart, vulva and paediatric neuroblastoma. All records from patient groups containing the target cancer were removed and the vocabulary was refitted before training; downstream analyses used the target-specific checkpoint. Lung and bronchus served only as a common-cancer control. The panel began with that control and three rare targets and was extended by six cancers under a protocol frozen before their target-specific runs; it is therefore a selected panel, not a random sample of unseen cancers. The primary estimand was leave-cancer-out pretrained minus random frozen. A descriptive comparison with the full-corpus encoder bundled corpus-size and vocabulary effects and was not a decision criterion. Full provenance and run artefacts are in Supplementary Methods S9.

#### 1.11.5 Conventional comparators

The conventional Cox comparator one-hot encoded the same 18 canonical diagnostic positions, with levels fitted on each target’s training split, and used ridge-penalised Efron Cox regression. The primary penalty was α = 1.0; α = 0.01, 0.1, 10 and 100 formed a prespecified sensitivity grid without validation-based selection. Budget-matched Cox used the identical sampled patients as the representation arm.

All Cox-based arms used time-invariant linear predictors. For neural heads, the target-specific Breslow baseline cumulative hazard [28] gave S(t|x) = exp{-H0(t) exp(eta(x))}; the scikit-survival comparator used its fitted survival function [29]. No Schoenfeld-residual test or covariate hazard-ratio interpretation was performed, and proportional-hazards diagnostics were not an eligibility gate. Harrell concordance assessed fixed-risk ranking, while horizon-specific metrics assessed prediction at 12, 36 and 60 months. Possible time-varying predictor effects remain a model-specification limitation. Input-identity checks and full configurations are in Supplementary Methods S7.

#### 1.11.6 End-to-end comparator

An end-to-end arm trained the architecture and Cox head jointly from random initialisation on the same patients, order, seed and horizons. To preserve the exact full-risk-set Efron objective without a mini-batch approximation, a two-pass procedure first computed full-cohort risks and per-example loss gradients without retaining activations, then recomputed chunked activations for back-propagation. CUDA state was restored so dropout masks matched across passes. This changed peak memory, not the loss or gradient, and was verified empirically. Supplementary Methods S7 gives the algorithm and checks.

### **1.12** Outcome metrics, uncertainty and interval classification

A shared evaluator required finite scalar risk and survival probabilities within [0,1] and non-increasing over time, then scored every arm identically. Harrell concordance was primary [30]; Uno concordance [31], cumulative/dynamic AUC [32], integrated Brier score [33], and grouped Kaplan–Meier calibration [34] were secondary. Calibration-in-the-large was observed minus mean predicted event probability. The binned absolute calibration error was the bin-size-weighted absolute difference between Kaplan–Meier observed and mean predicted event probability across 10 equal-count bins; it is not the smoothed individual-level integrated calibration index [35]. Horizons were 12, 36 and 60 months where both training and evaluation follow-up supported them.

Uncertainty used paired patient-group cluster bootstrap with 1,000 replicates for full-label analyses. Identical resampled patient groups were reused across arms within each contrast, and percentile limits were taken at 2.5% and 97.5%. Resumable runs partitioned bootstrap replicates only, never patient rows. Differences were contender minus named reference. Each Harrell-concordance difference was classified by an executable rule against zero and a locked, investigator-specified minimum important difference (MID) of 0.01; the MID is an analysis relevance threshold, not a validated clinical-utility threshold.

An interval entirely above the MID was positive and important; an interval above zero but spanning or remaining below the MID was positive with uncertain magnitude or positive but below importance, respectively. An interval entirely below zero was negative. Intervals spanning zero were underpowered when their half-width exceeded the MID and otherwise inconclusive with adequate precision. Classifications were target- and budget-specific. Intervals are nominal and unadjusted for multiplicity, so cross-target counts are descriptive and concordance was not pooled across cancers. Supplementary Methods S10–S12 gives formulas, censoring rules, bootstrap implementation and exact boundaries.

### **1.13** Governance of the held-out test partition

The test partition remained sealed until a checksummed release file confirmed that all nine validation results existed, prediction contracts and exact Cox checks passed, and checkpoint, source and split-manifest digests matched. The nine-target full-label leave-cancer-out analysis was the sealed first look. The 67-cancer Cox comparison at 256 labels, end-to-end comparison and rare-nine three-comparator analysis were protocolized exploratory extensions specified after that release. The test partition was therefore not globally untouched. Supplementary Methods S13 gives the release record and chronology.

### **1.14** Software and computation

PyTorch supported encoder training and probes; scikit-survival supported survival metrics and Cox models; DuckDB and PyArrow supported data construction. GPU training used an RTX 6000D, and large CPU analyses used a 96-core node. Checksums linked formal results to their code and inputs. An independent audit regenerated all 39 figure source tables, all 45 panels and all 46 numeric legend claims; every table and claim agreed, and 43 panels were pixel-identical. The released version-restricted code package includes the numbered workflow, coding gate, statistical methods and executable interval-classification rule, but no patient data, weights or predictions. Execution environments and the full audit are documented in Supplementary Methods S14–S15.

### **1.15** Ethics and data permissions

SEER Research Plus data were accessed under the applicable National Cancer Institute data-use agreement [36]. The analysis used existing registry data, involved no participant contact and performed no linkage to an external patient-level database. The project archive does not contain an institution-specific ethics-committee determination or consent waiver; the corresponding author must insert the verified institutional statement before submission. Data access alone is not treated here as evidence of ethics approval or exemption.

### **1.16** Analyses specified but not performed

Three planned analyses were not executed, and their absence is not a null result: temporal extrapolation, cancelled by decision and never evaluated; external validation, for which no independent cohort was available, leaving all evaluation internal to the SEER 17 registries on patient-disjoint splits; and registry holdout or domain-adversarial designs, made impossible by the absence of a registry identifier in the case-level export. What each omission does and does not permit is stated in the Discussion.

## **2.** Results

### **2.1** Cohort, temporal structure and input representation

Of 9,708,868 tumour records diagnosed in 2000–2023, 9,425,135 (97.08%) met eligibility criteria. Behaviour consistency and reporting source excluded 171,994 and 114,818 records, respectively; these counts are marginal, not sequential **(Figure 1A)**. Hash partitioning assigned 80.0%, 10.0% and 10.0% of records to training, validation and test with no patient crossover **(Figure 1B)**. Annual volume rose from 332,878 records in 2000 to 488,219 in 2023 **(Figure 1C)**; complete cohort routing and checkpoint relationships are in **Supplementary Figure S1**.

Follow-up maturity declined near the linkage cut-off. For diagnosis year 2000, status-known proportions at 12, 36 and 60 months were 0.997, 0.996 and 0.994; 60-month status-known fell to 0.374 in 2019, 36-month status-known to 0.291 in 2021, and all horizons to 0.123 in 2023 (**Figure 1D**). The 67 target cohorts ranged from 264 to 135,366 validation patients, a more than 500-fold range that constrains precision (**Figure 1E**; **Supplementary Table S1**). The raw and canonical views contained 22 and 18 field slots plus CLS, with 15 shared; only 10 of 23 inputs spanned all diagnosis years (**Figure 1F–G**; **Supplementary Table S2**).

### **2.2** Pretraining-objective diagnostic analysis

Pretraining ended at step 11,084 after 2.94 h; the terminal checkpoint alone was used downstream (**Figure 2A–C**). The sealed test contained 942,829 records, 1,885,658 raw and canonical sequences and 837,364 patient clusters. Of 33,941,844 masked targets, 33,941,788 were analysable and 56 unknown-value targets were excluded by rule. Primary NLL favoured the frequency baseline: 3.4063 versus 1.9215, a model-minus-baseline difference of +1.4848 (95% CI +1.4817 to +1.4876), so the prespecified reference point was not met (**Figure 2D**).

**Figure 2.**
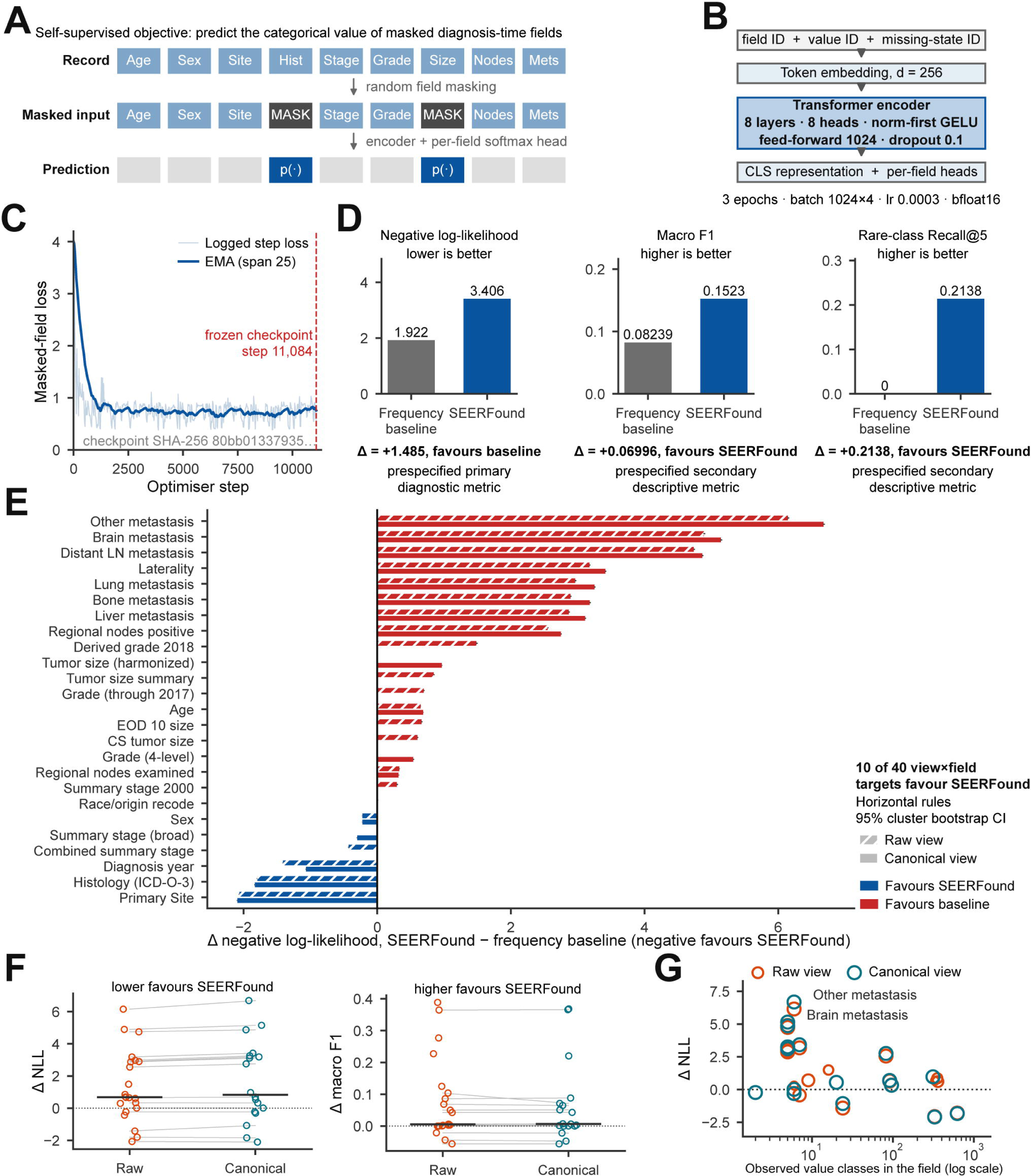
Pretraining objective, optimisation, and diagnostic evaluation. A, Self-supervised masked-field prediction task. B, Transformer encoder and field-specific prediction heads. C, Training loss and exponential moving average; the released checkpoint was frozen at step 11,084. D, Diagnostic evaluation of the pretraining objective on the test split, against the empirical-frequency baseline. Negative log-likelihood (NLL; lower is better), the prespecified primary diagnostic metric, favoured the baseline (SEERFound minus baseline, +1.4848; 95% CI, +1.4817 to +1.4876), whereas macro F1 (Δ = +0.0700) and rare-class Recall@5 (Δ = +0.2138) favoured SEERFound and are reported as prespecified secondary descriptive metrics. This analysis characterises the pretraining objective and does not condition the representation-structure or downstream-transfer analyses. E, Field-level NLL differences; negative values favour SEERFound, hatching denotes the raw view, and horizontal rules show 95% cluster-bootstrap CIs. F, Raw- and canonical-view differences in NLL and macro F1; grey lines link shared fields. G, Association between field cardinality and NLL difference; circle size represents the number of eligible masked targets. CI, confidence interval; NLL, negative log-likelihood.

The descriptive secondary metrics favoured the encoder: macro F1 was 0.1523 versus 0.0824 (difference +0.0700, 95% CI +0.0698 to +0.0704) and rare-class Recall@5 was 0.2138 versus 0.0000 (+0.2138, +0.2083 to +0.2198). Validation showed the same directions.

At field level, 10 of 40 view-by-field targets favoured the encoder on NLL and 30 favoured the baseline (**Figure 2E**). The largest adverse differences were for Other metastasis in canonical (+6.68) and raw (+6.15) views; overall raw- and canonical-view differences were +1.3613 (+1.3584 to +1.3640) and +1.6357 (+1.6323 to +1.6388), respectively (**Figure 2F**). The excess was concentrated in a minority of low-cardinality fields (**Figure 2G**). This localises reconstruction loss but does not establish probability miscalibration. Complete estimates and concentration analyses are in **Supplementary Table S3** and **Supplementary Figure S2**.

### **2.3** Representation structure and stability

Both branches used validation data only (**Figure 3A**). Across 10,000 patient groups, pretrained versus random Recall@1 was 0.9572 versus 0.5194, Recall@5 0.9886 versus 0.7374 and MRR 0.9713 versus 0.6206 (**Figure 3B**). Paired differences were +0.4378 (95% CI +0.4298 to +0.4459) for Recall@1 and +0.3507 (+0.3441 to +0.3574) for MRR, meeting the joint reference point (**Figure 3C**). Matched-pair separation was +2.21 standard deviations for the pretrained encoder versus +1.43 for random (**Figure 3D**). Mean ranks were 1.06 and 1.51 in the two pretrained retrieval directions, compared with 9.46 and 19.81 for random, whose errors were long-tailed (**Figure 3E**).

**Figure 3.**
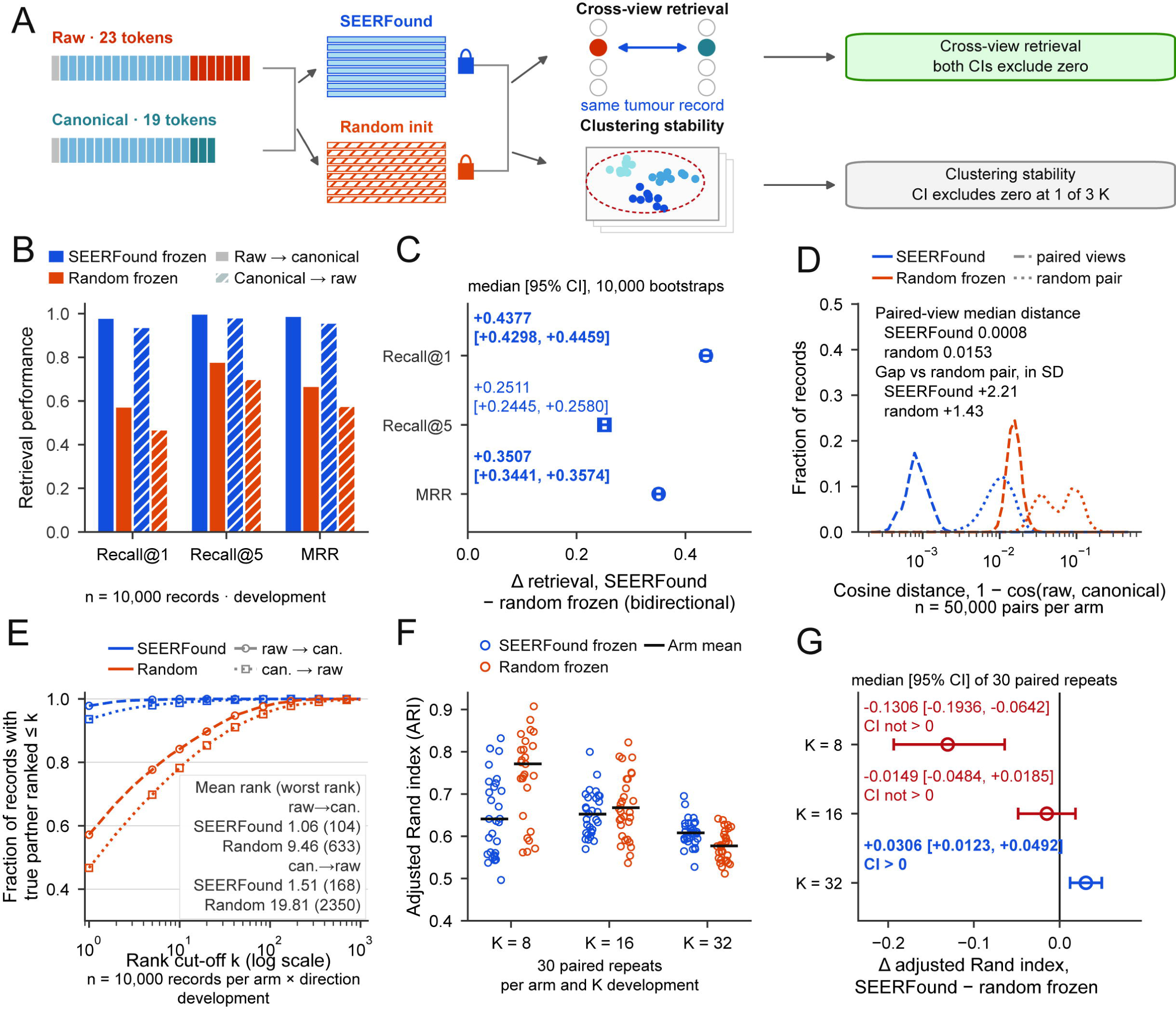
Cross-view representation alignment and clustering stability (development domain) A, Design and prespecified reference points. Frozen SEERFound and architecture-identical randomly initialised encoders were compared on two branches, cross-view retrieval and clustering stability; the branches are reported separately and neither is conditional on the other. B, Recall@1, Recall@5, and mean reciprocal rank (MRR) for raw-to-canonical and canonical-to-raw retrieval in 10,000 development records. C, Median paired retrieval differences between SEERFound and the random-frozen control, with 95% bootstrap CIs from 10,000 resamples. D, Cosine-distance distributions for matched raw–canonical records and randomly paired records (50,000 pairs per arm; exploratory). E, Fraction of true paired records recovered across rank cut-offs. F, Adjusted Rand index (ARI) across 30 paired repeats at K = 8, 16, and 32. G, Median paired ARI differences with 95% CIs; the lower limit was above zero at only one of three K values, short of the prespecified two, so this branch does not support a claim of more reproducible cluster structure. ARI, adjusted Rand index; MRR, mean reciprocal rank.

Clustering stability did not meet its reference point. Mean pretrained versus random ARI was 0.641 versus 0.771 at K = 8, 0.652 versus 0.668 at K = 16 and 0.608 versus 0.577 at K = 32. Paired differences were −0.1306 (−0.1936 to −0.0642), −0.0149 (−0.0484 to +0.0185) and +0.0306 (+0.0123 to +0.0492), respectively (**Figure 3F–G**). Only one of three lower limits exceeded zero, short of the required two; at K = 8 the interval lay entirely below zero.

### **2.4** Survival transfer in cancers represented in the pretraining corpus

Random frozen, end-to-end scratch and penalised Cox comparators address distinct boundaries of representation value (**Figure 4A**). Against the random frozen encoder, all 402 validation target-by-budget point estimates across 67 cancers were positive. Nominal intervals excluded zero in 29, 49, 64, 63, 62 and 60 of 67 cancers at 32, 64, 128, 256, 512 and 1,024 labels, respectively. Twenty-five cancers excluded zero at every budget and 49 at five or more; other male genital organs and paediatric neuroblastoma remained underpowered at all budgets. Differences were non-increasing from 128 to 1,024 labels in 61 cancers (**Figure 4B**; **Supplementary Table S4**, **Supplementary Figure S3**).

**Figure 4.**
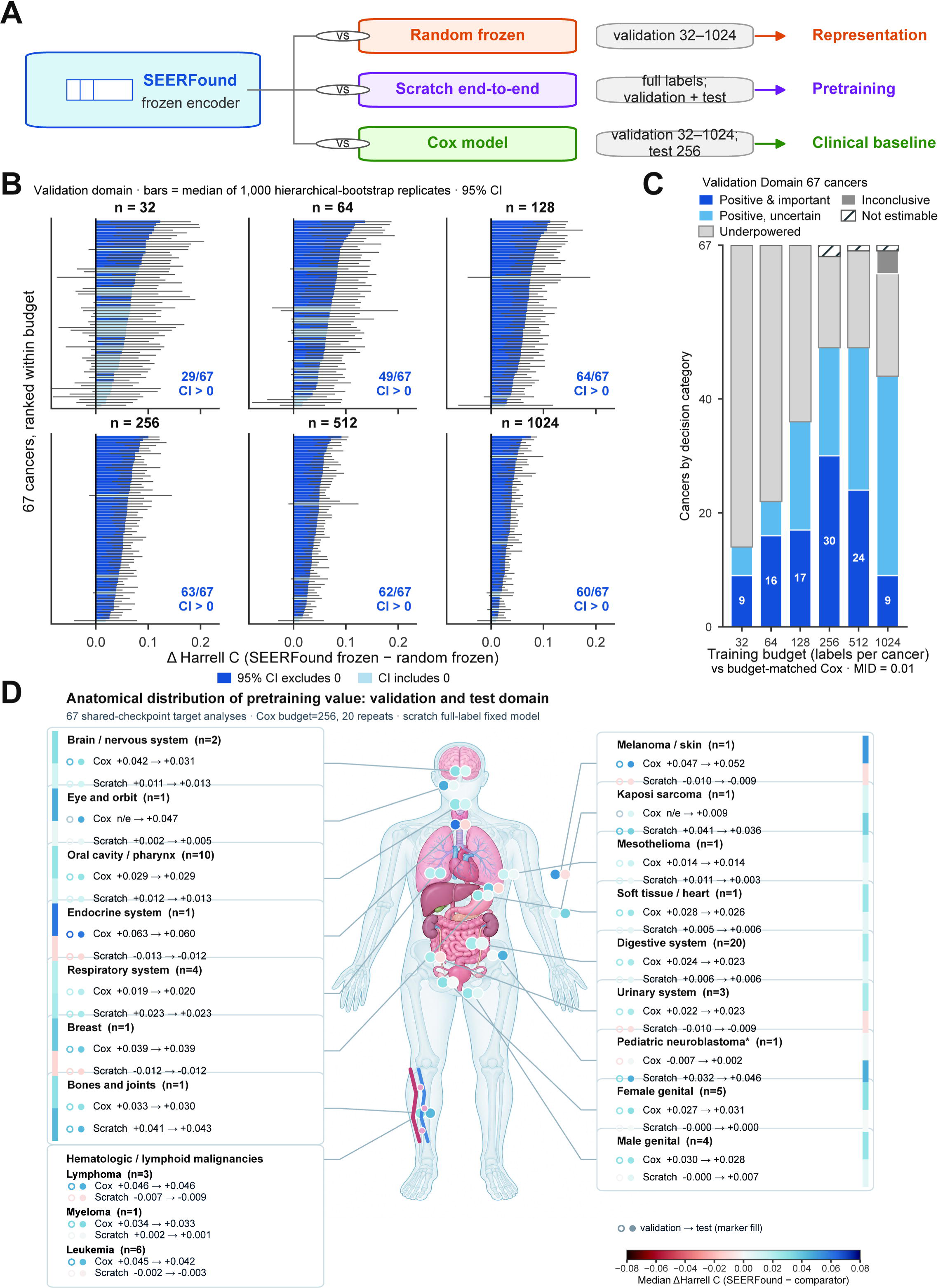
Boundary of pretraining value across comparators and evaluation domains. A, Comparator map. The random-frozen encoder tests representation value, end-to-end training from scratch tests the contribution of pretraining, and penalised Cox provides a clinical-feature baseline; these contrasts estimate different quantities. B, Validation-domain differences in Harrell C between frozen SEERFound and the random-frozen control across 67 cancers and label budgets of 32–1,024 patients. Points are medians of 1,000 hierarchical-bootstrap replicates and bars are nominal 95% CIs. C, Decision categories for SEERFound versus budget-matched Cox using a minimum important difference (MID) of 0.01; hatching denotes non-estimable comparisons. D, Cancer-group summaries for validation (open markers) and test (filled markers) against Cox and scratch comparators. These shared-checkpoint analyses are descriptive and are not strict LCO evaluations. CI, confidence interval; LCO, leave-cancer-out; MID, minimum important difference; n/e, not estimable.

Against budget-matched Cox, positive-and-important classifications across the six budgets were 9, 16, 17, 30, 24 and 9 of 67 cancers; underpowered classifications were 53, 45, 31, 16, 17 and 18 (**Figure 4C**). Positive-with-uncertain-magnitude classifications rose from 5 to 35, and four adequately precise inconclusive cells appeared only at 1,024 labels. At small budgets most comparisons were imprecise; by 256 labels positive-and-important classifications peaked, then shifted toward unresolved magnitude. The pattern indicates narrowing differences with more labels, not that residual differences were necessarily smaller than the MID.

At 256 labels in the protocolized test extension, all 67 point estimates favoured the frozen pretrained representation. The median difference was +0.0283 (range +0.0015 to +0.0752); 54 nominal intervals excluded zero and none lay entirely below it, yielding 26 positive and important, 28 positive with uncertain magnitude and 13 underpowered classifications. The largest differences included nodal Hodgkin lymphoma at +0.0752 (+0.0559 to +0.0921) and thyroid at +0.0601 (+0.0437 to +0.0738), while paediatric neuroblastoma was smallest at +0.0015 (−0.0488 to +0.0565) and underpowered. Absolute pretrained concordance ranged from 0.564 to 0.779. The 67 analyses represented 885,041 summed target-specific test memberships and 459,093 event memberships, not unique patients (**Supplementary Table S5**). Lung and bronchus, prostate and breast re-runs under the unified configuration yielded +0.0169, +0.0346 and +0.0386, respectively, with intervals above zero.

Against end-to-end training, 27 intervals favoured pretraining, 21 favoured scratch and 19 crossed zero. Advantages for pretraining included ureter (+0.048), peritoneum (+0.042) and paediatric neuroblastoma (+0.040); scratch was favoured in several large cohorts, including breast (−0.0109, −0.0118 to −0.0101) and lung and bronchus (−0.0076, −0.0082 to −0.0069). This cohort-size association is unadjusted and descriptive, and no mechanism is inferred (**Supplementary Table S5**). Corpus uteri, prostate and breast required removal of 22, 76 and 129 patients, respectively, from weighted secondary metrics at the unsupported 287-month endpoint; primary Harrell concordance retained the full cohorts.

Group medians across 19 anatomical or haematologic categories summarise both test and validation comparisons but do not replace target-level intervals (**Figure 4D**). Two Cox validation targets were not estimable, and breast end-to-end point estimates lacked completed intervals; these remain missing rather than being set to zero (**Supplementary Figure S4**).

### **2.5** Transfer to cancers absent from the pretraining corpus

Each of nine rare cancers was excluded from its own pretraining run, then evaluated once on sealed-test data with identical frozen Cox probes (**Figure 5A**). All nine pretrained-minus-random Harrell-concordance differences were positive with lower 95% limits above zero, ranging from +0.0034 (+0.0014 to +0.0052) for chronic lymphocytic leukaemia to +0.0368 (+0.0111 to +0.0645) for paediatric neuroblastoma (**Figure 5B**; **Supplementary Table S6**). Intermediate effects included nodal Hodgkin lymphoma +0.0245 (+0.0160 to +0.0320), soft tissue including heart +0.0199 (+0.0146 to +0.0250), mesothelioma +0.0166 (+0.0079 to +0.0254), salivary gland +0.0164 (+0.0098 to +0.0240), vulva +0.0149 (+0.0083 to +0.0213), acute myeloid leukaemia +0.0112 (+0.0082 to +0.0143) and myeloma +0.0079 (+0.0055 to +0.0104).

**Figure 5.**
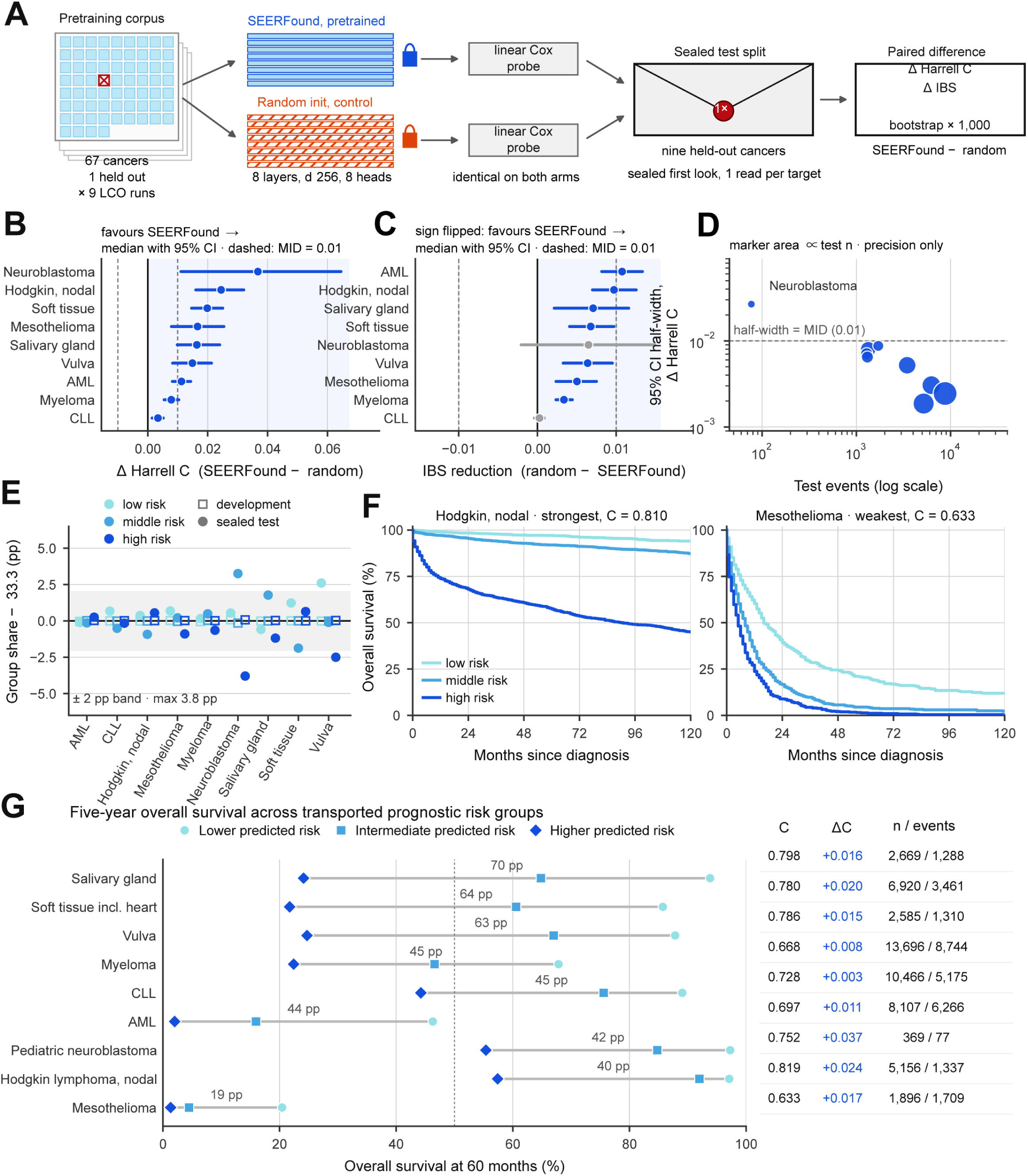
Sealed-test transfer under leave-cancer-out pretraining. A, LCO design for nine rare cancers. Each target cancer was excluded from pretraining; pretrained and random-frozen encoders then received identical linear Cox probes and were evaluated once on the sealed test split. B, Test-set difference in Harrell C (SEERFound minus random) with 95% bootstrap CIs; all nine estimates were positive. C, Reduction in integrated Brier score (IBS; random minus SEERFound), with positive values favouring SEERFound. D, Test-event count versus CI half-width; marker area is proportional to test cohort size. E, Difference between test risk-group proportions and the one-third target, using tertile thresholds fixed in development data. F, Kaplan–Meier curves for the validation-selected strongest and weakest target cancers. G, Five-year overall survival across transported low-, intermediate-, and high-risk groups for all nine cancers. Risk-group analyses show prognostic separation and do not establish treatment benefit or clinical utility. CI, confidence interval; IBS, integrated Brier score; KM, Kaplan–Meier; LCO, leave-cancer-out; MID, minimum important difference.

Seven point estimates reached the MID of 0.01, but a point estimate crossing the threshold is not the same as the interval-based threshold being met. Under the stricter rule, paediatric neuroblastoma, nodal Hodgkin lymphoma and soft tissue including heart were positive and important; five cancers were positive with unresolved magnitude and chronic lymphocytic leukaemia was positive but below importance. None was negative, underpowered or inconclusive. Absolute pretrained concordance ranged from 0.633 in mesothelioma to 0.819 in nodal Hodgkin lymphoma.

Integrated Brier score improved with a lower limit above zero in seven of nine targets; paediatric neuroblastoma (+0.0065, −0.0021 to +0.0149) and chronic lymphocytic leukaemia (+0.0003, −0.0004 to +0.0009) crossed zero (**Figure 5C**). Interval precision generally increased with event count; paediatric neuroblastoma, with 77 test events, was the sole target whose half-width exceeded 0.01 (**Figure 5D**). Thus discrimination and calibration did not move together in every target.

Validation-derived risk tertiles transported to test within 3.8 percentage points of equal thirds (**Figure 5E**). Kaplan–Meier displays were pre-selected by validation concordance, using nodal Hodgkin lymphoma (0.810) and mesothelioma (0.633) (**Figure 5F**). Across all nine targets, low- versus high-risk 60-month OS differences ranged from 19.1 percentage points in mesothelioma to 69.7 in salivary gland cancer (**Figure 5G**; **Supplementary Table S7**, **Supplementary Figure S5**). These strata define no treatment threshold, treatment effect or clinical utility. Full horizon-specific calibration is in **Supplementary Table S8** and **Supplementary Figure S6**.

### **2.6** Low-label performance in rare cancers and its boundary

Four analyses locate the budget-dependent boundary against conventional modelling: paired few-shot Cox, full-data Cox, Cox-penalty sensitivity and a fixed-budget test extension (**Figure 6A**).

**Figure 6.**
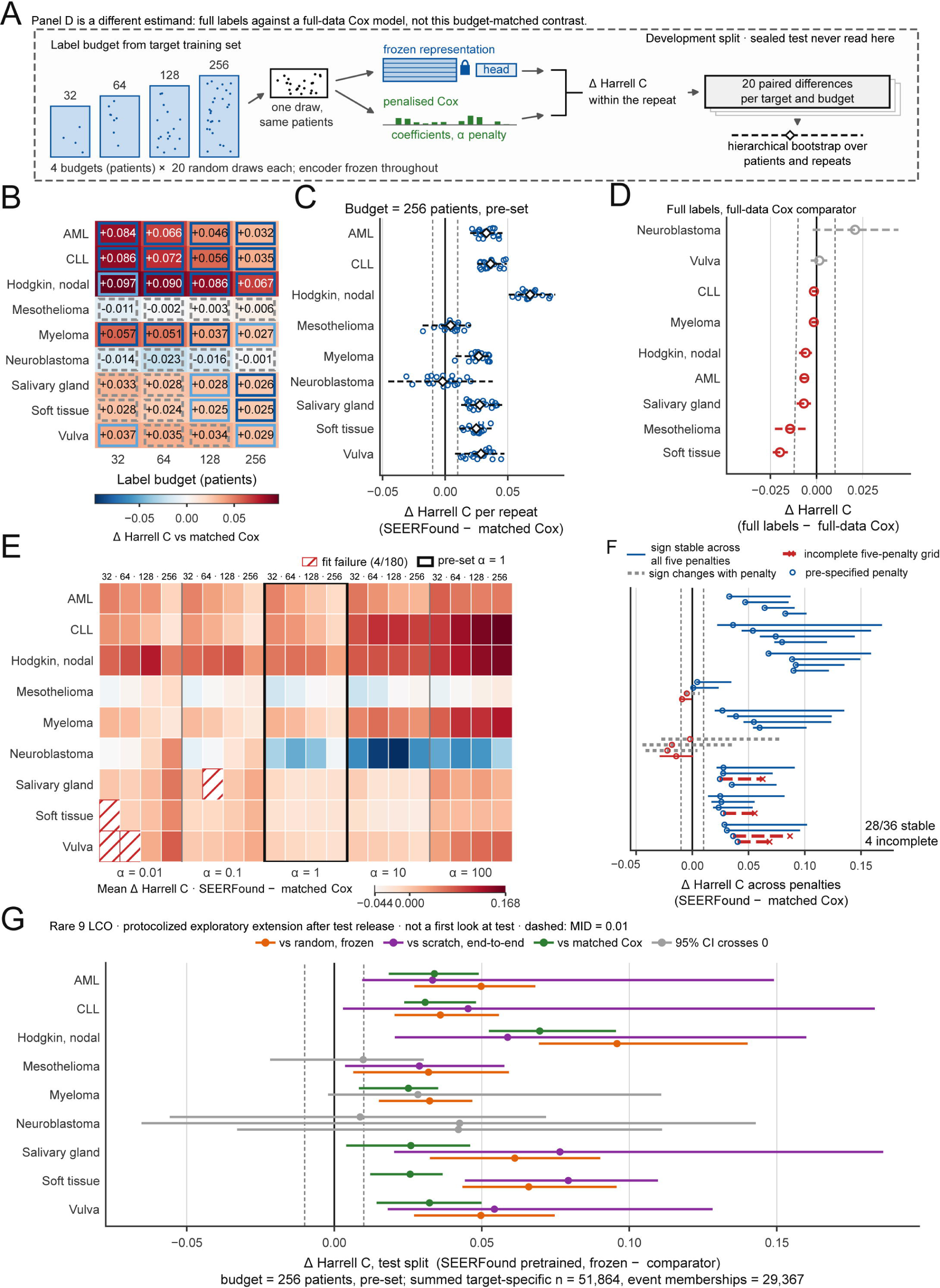
Low-label application in rare cancers and robustness boundary. A, Development-domain paired few-shot protocol: frozen SEERFound and penalised Cox used the same sampled patients at label budgets of 32, 64, 128, and 256, with 20 repeats per target and budget. B, Median differences in Harrell C versus budget-matched Cox; cell colour shows effect size and borders show interval-based classification. C, Repeat-level differences at the pre-set 256-patient budget; diamonds and dashed bars show hierarchical-bootstrap medians and 95% CIs. D, Full-label comparison with full-data Cox, a separate estimand from the budget-matched analysis. E, Sensitivity to Cox penalty α = 0.01–100; the black box marks the pre-set α = 1 and hatching denotes fit failure. F, Sign stability across penalties; 28 of 36 complete target–budget combinations retained the same sign. G, Exploratory post-release test extension at 256 patients against random-frozen, scratch, and matched-Cox comparators; grey intervals cross zero. CI, confidence interval; LCO, leave-cancer-out; MID, minimum important difference.

#### 2.6.1 Few-shot advantage against budget-matched Cox models

All 36 rare-cancer target-by-budget cells at 32–256 labels completed 20 repeats. Against Cox fitted to the same patients, 16 cells were positive and important, 6 positive with uncertain magnitude and 14 underpowered; none was negative (**Figure 6B**). Among seven targets with positive effects, the advantage generally narrowed with budget: acute myeloid leukaemia fell from +0.084 at 32 labels to +0.032 at 256, and nodal Hodgkin lymphoma from +0.097 to +0.067. Five of seven declined strictly; salivary gland and soft tissue had local rebounds but ended below their 32-label effects. Mesothelioma was −0.011 at 32 labels, and paediatric neuroblastoma −0.014 and −0.023 at 32 and 64; both were underpowered throughout and had the widest repeat dispersion at 256 labels (**Figure 6C**; **Supplementary Table S9**, **Supplementary Figure S7**).

#### 2.6.2 Boundary of the advantage at full label availability

With the full target training set, the direction reversed: seven of nine differences favoured full-data Cox with upper limits below zero, ranging from −0.0199 (−0.0237 to −0.0156) for soft tissue including heart to −0.0014 (−0.0025 to −0.0004) for chronic lymphocytic leukaemia. Vulva (+0.0014, −0.0032 to +0.0058) and paediatric neuroblastoma (+0.0208, −0.0021 to +0.0443) crossed zero. No lower limit exceeded zero (**Figure 6D**; **Supplementary Table S10**). This full-label estimand is distinct from the budget-matched comparison.

#### 2.6.3 Sensitivity of the low-label comparison to the Cox penalty

Across α = 0.01–100, direction was stable in 28 of 36 target-by-budget grids; four complete grids changed sign and four were incomplete (**Figure 6E–F**). Of 180 cells, 176 were estimable, and the α = 1 control reproduced all 720 frozen repeat differences within 10□¹². Sign changes occurred for mesothelioma at 64 labels and paediatric neuroblastoma at 64, 128 and 256. Non-estimable fits were salivary gland at α = 0.1 and 64 labels, soft tissue at α = 0.01 and 32 labels, and vulva at α = 0.01 and 32 or 64 labels. No validation result selected α and test data were not read. Penalty sensitivity occurred in the two targets underpowered throughout, but no common cause is inferred (**Supplementary Table S11**). The shared-checkpoint grid in **Supplementary Figure S8** estimates a different contrast.

#### 2.6.4 Fixed-budget test extension across three comparators

At 256 labels in the protocolized test extension, intervals favoured the pretrained leave-cancer-out arm in 8 of 9 targets versus random frozen and in 7 of 9 versus both end-to-end and budget-matched Cox; no interval in any contrast lay entirely below zero (**Figure 6G**). Against random, effects ranged from +0.036 (+0.021 to +0.055) in chronic lymphocytic leukaemia to +0.096 (+0.070 to +0.140) in nodal Hodgkin lymphoma; paediatric neuroblastoma crossed zero. Against end-to-end, the largest effect was soft tissue at +0.079 (+0.045 to +0.109), while myeloma and paediatric neuroblastoma crossed zero. Against Cox, nodal Hodgkin lymphoma reached +0.070 (+0.053 to +0.095), while mesothelioma and paediatric neuroblastoma crossed zero. The analyses contained 51,864 summed target-specific test memberships and 29,367 event memberships, not unique patients or a pooled estimand. Per-target estimates and denominators are in **Supplementary Table S12**.

## Discussion

This study addressed three separate questions about a masked-pretrained encoder for harmonised pan-cancer registry records. First, the pretraining-objective diagnostics were mixed: negative log-likelihood favoured the field-frequency baseline by +1.4848 (95% CI +1.4817 to +1.4876), whereas macro F1 and rare-class Recall@5 favoured the encoder. Second, the pretrained representation improved cross-view retrieval over an architecture-identical random encoder by +0.4378 in Recall@1 and +0.3507 in mean reciprocal rank, but clustering stability exceeded random at only one of three cluster counts. Third, downstream transfer was positive under limited labels but did not replace full-data conventional modelling. At 256 labels, all 67 cancers favoured the frozen pretrained representation over budget-matched Cox models, with a median concordance difference of +0.0283. All nine cancers excluded from pretraining favoured the pretrained over the random frozen encoder, with nominal intervals excluding zero and differences from +0.0034 to +0.0368; however, full-data Cox models were favoured in seven of those nine cancers. The evidence therefore supports a bounded label-scarce use case, not a single overall claim of model superiority.

The upstream and downstream results need not agree because they measure different properties. Thirty of 40 view-by-field targets favoured the frequency baseline on NLL, while macro F1 and rare-class Recall@5 favoured the encoder by +0.0700 and +0.2138. This pattern does not establish probability miscalibration or explain the survival results; neither question was tested. It instead reinforces a narrower methodological point: reconstruction performance should be treated as a diagnostic and should not substitute for direct evaluation of downstream transfer [16,37,38].

The closest prior studies differ mainly in what is withheld, whether survival labels enter pretraining, and which comparator defines benefit (Supplementary Table S13). Structured-data foundation models have generally evaluated transfer within the same source population [10,11,12,14], and event-stream pretraining has shown label efficiency and cross-institution transfer without holding out an entire disease [13]. In molecular cohorts, pan-cancer transfer, meta-learning and mixture-of-experts approaches have improved survival prediction under limited or absent target labels, but pretraining or joint training used survival outcomes [18,19,20]. Registry studies have used supervised multitask learning across related SEER cancers [21] or transferred supervised models from SEER to an external colorectal cohort [39]. The present contribution is narrower: outcome-agnostic pretraining on cross-era verified registry fields, complete exclusion of the target cancer with vocabulary refitting, and comparison with both budget-matched and full-data conventional models. The architecture-identical random frozen encoder isolates the contribution of pretraining more closely than an unrelated baseline, but it is not a null model; it remains a random non-linear feature map. Leave-cancer-out pretraining likewise tests disease-level transfer rather than patient-level holdout, although its comparison with the full-corpus encoder cannot separate the effects of a smaller corpus from those of a changed vocabulary.

The boundary of the benefit was consistent across comparisons. Among the nine unseen cancers, 16 of 36 target-by-budget cells favoured the pretrained representation by at least the prespecified relevance threshold, 6 were positive with unresolved magnitude and 14 were underpowered; the advantage generally narrowed from 32 to 256 labels. Full-data Cox models reversed the direction in seven of nine cancers. Across 67 targets, pretraining won the three-arm comparison in 27 and end-to-end training in 21, with a descriptive association between direction and cohort size. These findings accord with evidence that classical models remain competitive on tabular data [40,41] and that deep survival models do not uniformly outperform Cox regression [42], but the cross-cancer association cannot isolate the causal effect of label availability. The practical interpretation is therefore limited to settings with no more than a few hundred labelled patients.

Effect size also constrains interpretation. Concordance differences of +0.003 to +0.037 are small, and concordance can be insensitive to clinically relevant changes in individual risk estimates [43]. The prespecified 0.01 threshold is an analysis rule, not a validated threshold for clinical utility; net benefit was not assessed [44]. Integrated Brier score differences favoured pretraining in seven of nine unseen cancers but crossed zero in paediatric neuroblastoma and chronic lymphocytic leukaemia, so discrimination and probabilistic accuracy were not uniformly aligned [45]. Likewise, the 60-month Kaplan–Meier separations of 19.1 to 69.7 percentage points describe prognostic strata under a frozen score, not treatment thresholds, treatment effects or clinical utility.

The rare-cancer setting is both the most relevant and the least certain. Rare cancers collectively account for a substantial disease burden, but individual cancers often provide too few events for well-powered cancer-specific models [3,4,5]. The nine-cancer panel broadened haematological and rare-solid representation and was frozen before leave-cancer-out training, but it was selected rather than randomly sampled; the proportion benefiting cannot be generalised beyond it. Paediatric neuroblastoma, with 369 test patients and 77 events, had the widest interval, changed sign across the Cox penalty grid at three of four budgets and remained underpowered; mesothelioma was similarly uncertain at the smallest budgets. Small event counts are compatible with these patterns, but model or data instability cannot be excluded.

The main strengths were the fail-closed coding-verification gate, repeated conservation and leakage checks, patient-group splitting with verified zero crossover, unique index tumours and a shared evaluator for all model arms. These features address common sources of leakage and irreproducibility in prediction-model studies [46,47]. A release gate governed the test partition, interval classifications were executable, reporting followed STROBE, RECORD and TRIPOD+AI [22,23,24], and an independent audit reproduced the figure source tables and rendered panels from frozen artefacts.

Several limitations remain. All results are internal to patient-disjoint splits from the SEER 17 registries; no external cohort was available, and independent validation is required before claims about generalisability [48]. Registry-level holdout and domain-adversarial evaluation were not possible because the case-level export lacks a registry identifier. Temporal extrapolation was not evaluated, so no claim is made about future diagnosis years. Inputs were limited to 23 verified diagnosis-time fields. Fine-grained AJCC and EOD T, N and M categories, site-specific factors and biomarkers lacked verifiable era-consistent coding; chemotherapy and hormone-therapy data are not released, and recorded first-course treatment has only moderate sensitivity against claims-based ascertainment [49]. Treatment, comorbidity, performance status and recurrence were therefore absent, and the outcome was overall rather than cancer-specific survival. Cox-based arms assumed a time-invariant linear predictor; Schoenfeld-residual testing was not performed, and no covariate hazard ratio was interpreted. Intervals were nominal and unadjusted for multiplicity, no family-wide null hypothesis was tested, and the 67 targets included nine anatomically nested colorectal subsites, making cross-target counts descriptive. Finally, the study was not registered with a third-party timestamp. Protocol ordering rests on checksummed project records; the reconstruction plan was frozen after other analyses had produced results but before any reconstruction prediction existed. Only the nine-target leave-cancer-out analysis was a sealed first look; the other test-domain analyses were protocolized exploratory extensions specified after that release.

The route to clinical use remains substantial, particularly in rare cancers where prognostic evidence is sparse [3,4]. It begins with external validation across registries and health systems with different coding practices [48], followed by calibration assessment [45], decision-curve analysis at clinically relevant thresholds [43,44] and prospective evaluation in a care pathway. Staging detail, site-specific factors and biomarkers excluded by the coding gate should also be introduced individually to determine whether the low-label advantage persists when conventional comparators receive richer inputs. None of these steps was completed here.

In conclusion, masked self-supervised pretraining produced a view-invariant registry representation with transferable prognostic signal under limited labels, including in nine cancers excluded from pretraining. The upstream diagnostics were mixed, and full-data Cox models outperformed the pretrained approach in seven of those nine cancers. The study therefore identifies a label-scarce setting in which the representation merits external evaluation, while providing no evidence of clinical utility, treatment benefit, deployment readiness or generalisation beyond the cancers, coding eras and settings studied. When sufficient labelled data are available, conventional survival modelling remains the default.

## DATA AVAILABILITY STATEMENT

Funding: The study was approved by the committee of Zhejiang University School of Medicine, Sir Run Run Shaw Hospital. This work was supported by the Natural Science Foundation of China (No: 82402697and 82472351), Natural Science Foundation of Zhejiang Province (ZCLQN25H2001, and LQN25H160028), Fujian Provincial Science and Technology Innovation Joint Fund Project (2024Y9698), Natural Science Foundation of Xiamen (3502Z202471063) and Xiamen Qinglu Talent Start-up Funding (K2024-02).

## Data Availability

All data produced in the present work are contained in the manuscript and available online at https://seer.cancer.gov/data/

## Supplementary figure legends, Figures S1–S8

Supplementary Figure S1. Cohort routing, checkpoint relationships and evidence domains

A, Routing of registry records, patient-hash splits, pretraining relationships and downstream analysis families. The source registry held 9,708,868 tumour records diagnosed in 2000–2023, of which 9,425,135 met the Phase 1 modelling eligibility criteria. Deterministic splitting by released patient-identifier group gave 7,767,009 training, 970,778 validation and 971,081 test records, from 6,885,414, 860,909 and 860,599 patient groups respectively; eligibility and split counts are marginal and are not sequential subtractions. The training split fitted either one shared base checkpoint, in which every target cancer was represented during pretraining, or nine target-specific strict leave-cancer-out (LCO) checkpoints, each omitting one target cancer. The shared checkpoint underlies Figures 2 and 3, the strict-LCO checkpoints Figures 5–6. Domain badges distinguish development or validation analyses, the original sealed test analyses and protocolized exploratory extensions specified after test release. B, Evidence-domain matrix giving the pretraining relationship, target-label budget, evaluation domain, encoder state, comparator family and primary question of each analysis family. Shared-checkpoint and strict-LCO results address different estimands and were not pooled. LCO, leave-cancer-out.

Supplementary Figure S2. Cross-metric discordance and concentration of field-level negative log-likelihood excess

Diagnostics across the 40 raw- or canonical-view field targets of Figure 2. A, Joint direction of the field-level differences between SEERFound and the frequency baseline in negative log-likelihood (NLL; lower is better) and macro F1 (higher is better). Orange circles denote raw-view and teal squares canonical-view targets; marker fill redundantly encodes Recall@5, with coloured markers favouring SEERFound, white markers not evaluable because the verified number of rare-cancer targets was zero, and grey markers tied. Ten targets favoured SEERFound on both metrics, 20 favoured the baseline on NLL but SEERFound on macro F1, eight favoured the baseline on both, and two lay on the macro-F1 tie line; none favoured SEERFound on NLL while favouring the baseline on macro F1. B, Pareto diagnostic over the 30 targets with positive ΔNLL, ordered by decreasing ΔNLL, with the black curve giving the cumulative share of total positive NLL excess. The seven largest adverse targets accounted for 50.7% of that excess and the 14 largest for 81.0%. C, Post-hoc descriptive exclusion sensitivity: the unweighted mean ΔNLL was +1.4848 across all 40 targets and first turned negative, at −0.0304, after removing the 15 largest adverse targets. This characterises concentration; it does not replace the full 40-target comparison or alter the prespecified primary and secondary metric roles. NLL, negative log-likelihood.

Supplementary Figure S3. Fixed-order cancer-level trajectories across label budgets

Validation-set differences in Harrell concordance across 67 cancer targets and six label budgets of 32–1,024 patients, from 20 paired training repeats per target-by-budget cell. One cancer order, set by the frozen-representation-minus-Cox difference at 256 labels, is held across both panels and all budgets. Purple labels and diamonds identify the nine targets later examined with strict-LCO checkpoints in Figures 5–6; all results here use the shared base checkpoint. Cell fill spans a common scale of −0.14 to +0.14. A, Versus the random frozen encoder, plotting the median of 1,000 paired hierarchical-bootstrap replicates with the 2.5th to 97.5th percentile interval. All 402 point differences were positive; a black inset border marks a nominal, multiplicity-unadjusted 95% lower bound above zero, in 29, 49, 64, 63, 62 and 60 of 67 targets from the smallest to the largest budget. B, Versus the budget-matched Cox model, plotting instead the mean of the 20 observed paired repeat differences with its hierarchical-bootstrap interval. Inset borders encode the frozen decision classification independently of cell colour: dark blue, positive and important; light blue, positive with uncertain magnitude; dashed grey, underpowered; dotted black, inconclusive with adequate precision. Hatched cells were not estimable and were not assigned a value of zero. LCO, leave-cancer-out.

Supplementary Figure S4. Target-level validation-to-test consistency across conventional and end-to-end comparators

A, Validation versus test differences in Harrell concordance for the frozen pretrained representation against a budget-matched Cox model at 256 labelled patients per cancer. Each circle is one cancer target and marker area is proportional to test events; the dashed diagonal marks exact validation-to-test agreement and the dotted lines no difference. Paired estimates were available for 65 targets; eye and orbit and Kaposi sarcoma were not estimable in validation because of Cox fitting failures and were not assigned a value of zero. Spearman correlation was 0.866, with 64 of 65 targets agreeing in direction. B, Corresponding full-label comparison with the end-to-end scratch model across 67 targets; Spearman correlation 0.946, with 63 of 67 agreeing in direction. Breast cancer had point estimates in both domains but no completed bootstrap interval and is marked with a cross. C, Target-level test-minus-validation differences for both comparators; open circles are targets, diamonds medians and bars interquartile ranges. D, Exploratory cross-cancer Spearman associations between the development-domain pretrained-minus-scratch difference and validation sample size, validation event count, event fraction and training sample size, with counts log10-transformed; circles are estimates and bars 95% intervals from 2,000 bootstrap replicates, n = 67 cancers. Panels A–C are protocolized exploratory extensions specified after the original sealed test release.

Supplementary Figure S5. Complete sealed-test Kaplan–Meier risk stratification across nine leave-cancer-out targets

Kaplan–Meier overall-survival curves for the nine cancer targets excluded from their respective strict-LCO pretraining runs. Risk-score tertile cut-points were fitted in the validation split and transported without refitting to the sealed test split. Light-blue circles, medium-blue squares and dark-blue diamonds at 24, 60 and 120 months identify the lower, intermediate and higher predicted-risk groups; the dotted vertical line marks 60 months. Each facet reports the sealed-test sample size, event count and absolute Harrell concordance of the frozen pretrained representation, and the table beneath gives numbers at risk in the lower (L), intermediate (M) and higher (H) risk groups at 0, 24, 60 and 120 months. Across targets, test sample sizes ranged from 369 to 13,696, event counts from 77 to 8,744 and concordance from 0.633 to 0.819. The displayed strata are prognostic risk groups and define no treatment threshold, treatment effect or clinical decision rule. Confidence bands were not available in the frozen publication source data. LCO, leave-cancer-out.

Supplementary Figure S6. Complete calibration diagnostics across nine leave-cancer-out targets

Calibration of the frozen pretrained representation in the sealed test split. A, Binned absolute calibration error at 12, 36 and 60 months for each of the nine strict-LCO targets, defined as the bin-size-weighted mean absolute difference between the Kaplan–Meier observed and the mean predicted event probability across ten equal-count bins of predicted event probability. This is a descriptive binned summary and is deliberately not the integrated calibration index (ICI), which averages the absolute difference between predicted risk and a smoothed individual-level calibration curve; the two are not interchangeable and no ICI was computed. Within each cell the first value is that error and the parenthesized value is calibration-in-the-large (CITL), the observed minus the mean predicted event probability. Across the 27 displayed cells, all with ten estimable bins, the error ranged from 0.006 to 0.039 and CITL from −0.016 to +0.024. B, Reliability curves for all nine targets using ten equal-count prediction bins at each horizon. Circles, squares and diamonds denote 12, 36 and 60 months and marker area is proportional to the number of records in the bin; the dashed diagonal indicates perfect calibration and all facets share 0%–100% axes. Only the frozen pretrained arm is shown, to preserve legibility of the 27 curves; both model arms are reported in Supplementary Table S8. Bin-level confidence bands were not available in the frozen source data. CITL, calibration-in-the-large; ICI, integrated calibration index.

Supplementary Figure S7. Repeat-level stability of rare-cancer low-label transfer against budget-matched Cox models

Repeat-level differences in Harrell concordance between the strict-LCO frozen pretrained representation and the budget-matched Cox model for nine rare-cancer targets at label budgets of 32, 64, 128 and 256 patients. Open circles show the 20 paired training-sample repeats per target-by-budget cell, 720 paired values across 36 summaries. Filled diamonds give the median paired difference and vertical bars the paired hierarchical-bootstrap 95% confidence interval from 1,000 replicates reported in the frozen Figure 6 analysis. The solid horizontal line denotes no difference and the dashed line the prespecified minimum important difference (MID) of 0.01. All facets share a common y-axis so that between-target dispersion is not obscured by target-specific scaling. These are development-domain strict-LCO low-label comparisons and should not be combined with the full-label sealed-test estimand of Figure 5. LCO, leave-cancer-out; MID, minimum important difference.

Supplementary Figure S8. Cox ridge-penalty sensitivity for the shared base-pretrained checkpoint

Sensitivity analysis for the pan-cancer shared-base-checkpoint estimand of Figure 4; it does not assess the strict-LCO estimand of Figure 6, including for cancer names appearing in both analyses. A, Mean paired difference in Harrell concordance between the frozen shared base-pretrained representation and a label-budget-matched ridge-penalised Cox model across 13 cancers, five fixed penalties (α = 0.01, 0.1, 1, 10 and 100) and four label budgets (32, 64, 128 and 256 patients, ordered left to right within each penalty block), giving 260 cells. Blue denotes a negative and red a positive difference; two non-estimable Cox fits are hatched rather than assigned a value of zero, and the heavy outline marks the prespecified α = 1 block. B, Penalty sensitivity within each cancer-by-budget cell: the horizontal segment spans the range of differences across estimable penalties and the open circle gives the difference at the prespecified α = 1. Solid blue segments denote a stable sign across all five penalties, dotted grey segments a sign change, red segments and crosses an incomplete five-penalty grid, and red open circles a negative difference at the prespecified penalty. Twenty-six of 52 cancer-by-budget cells had a stable sign across all five penalties and two had incomplete grids. LCO, leave-cancer-out.

