## Supplementary figures and images for "Pretrained transformers applied to population cancer registries improve survival prediction in label-scarce and previously unseen cancers"

### Supplementary Figure S1

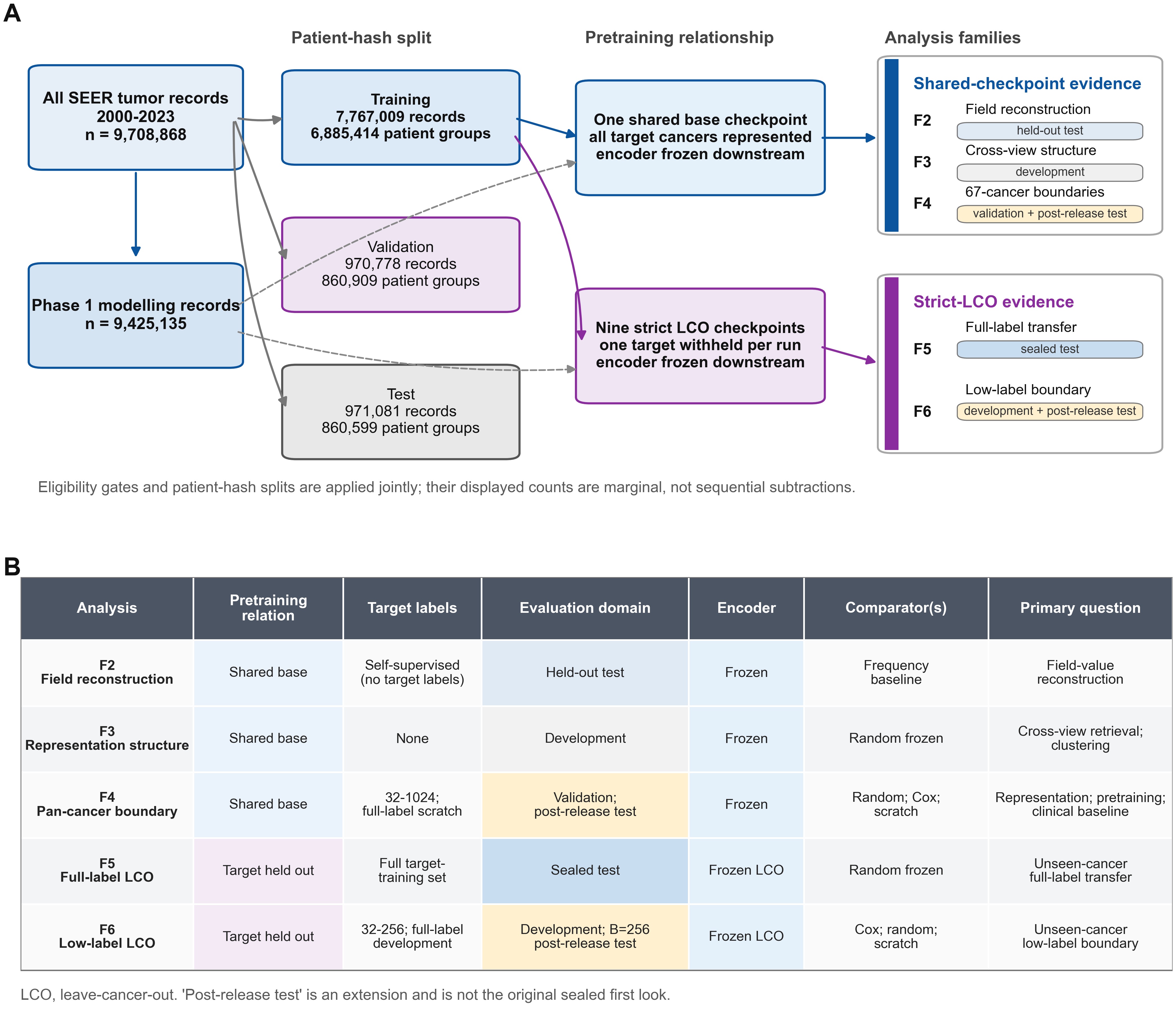

### Supplementary Figure S2

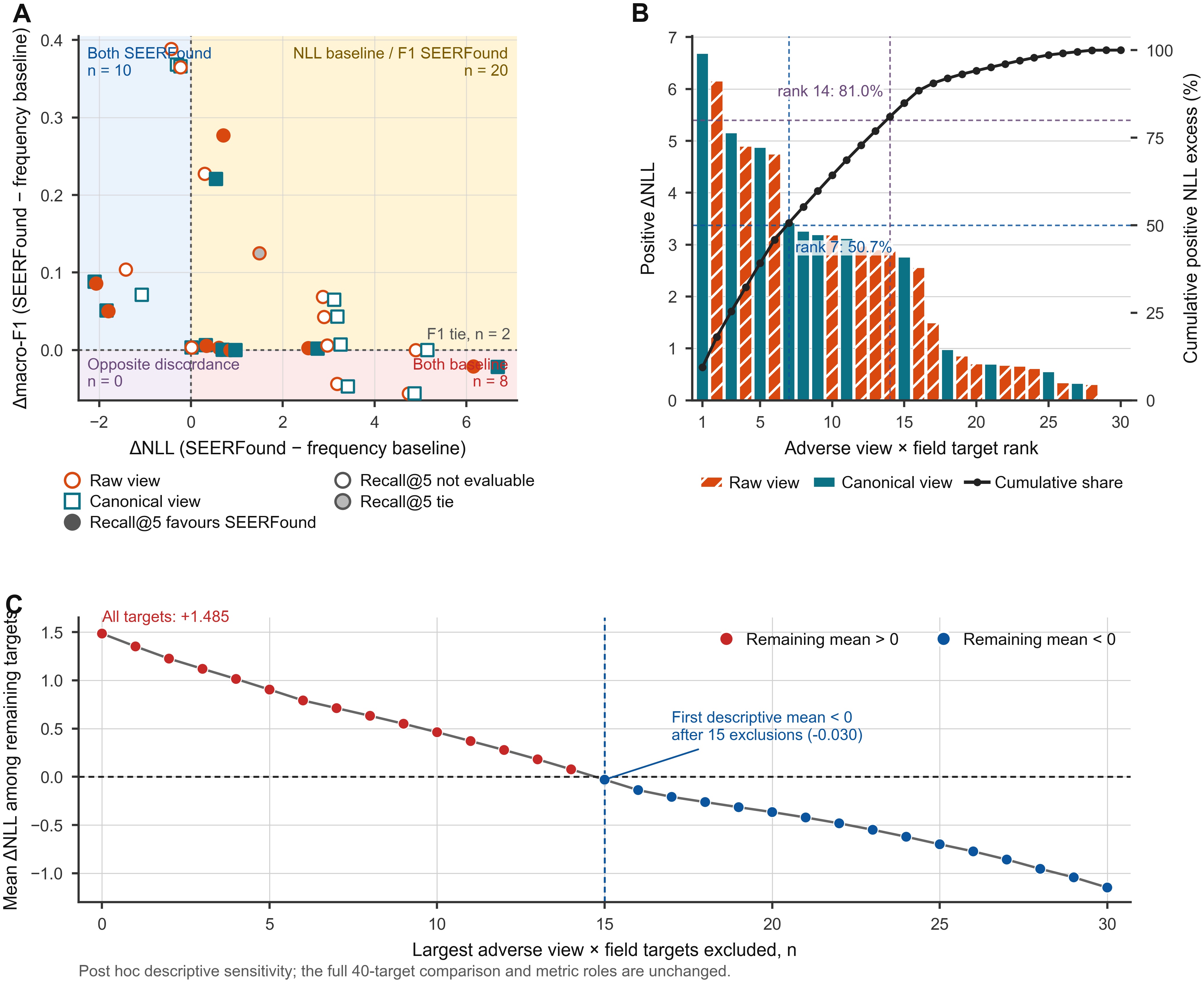

### Supplementary Figure S3

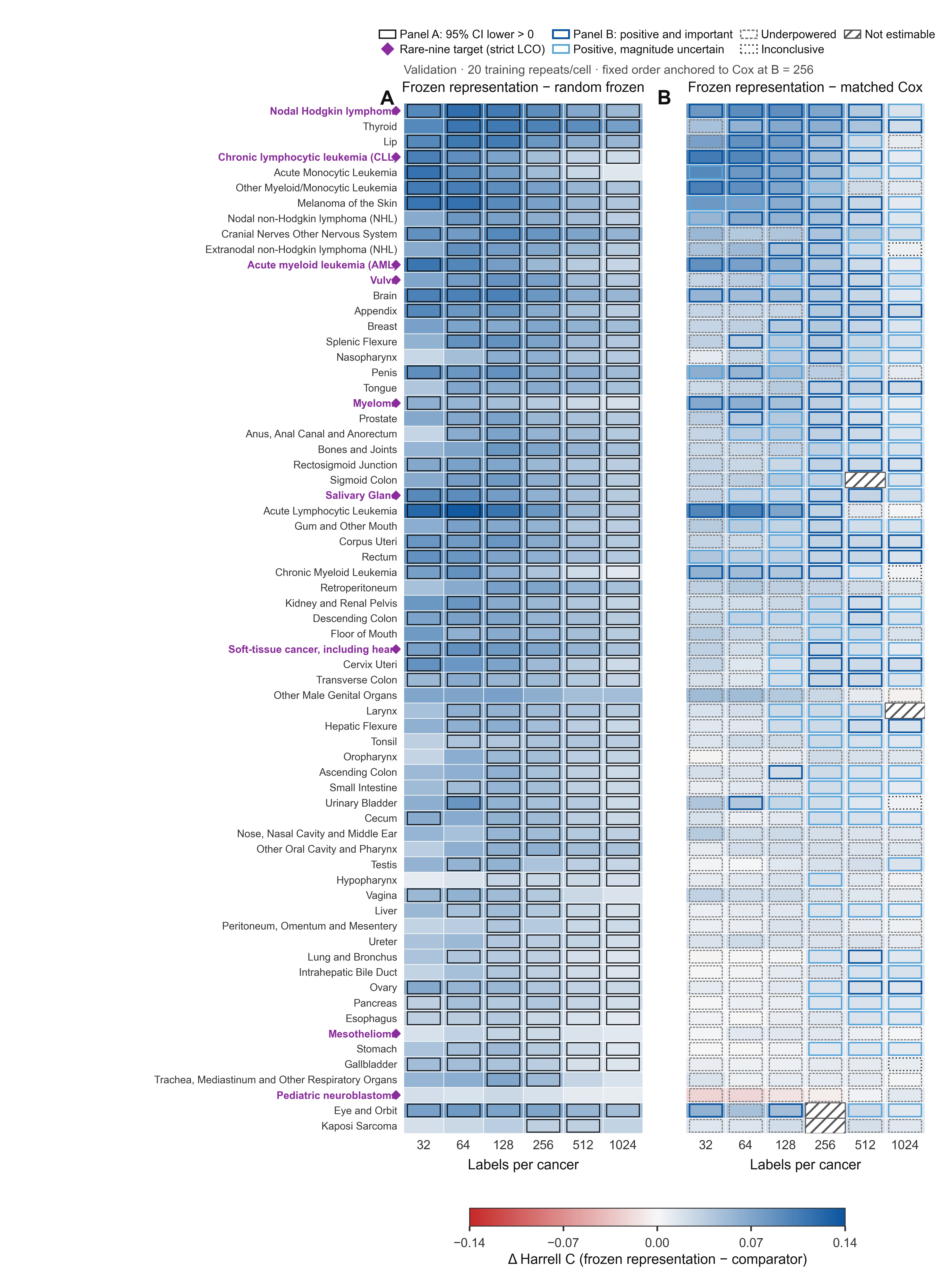

### Supplementary Figure S4

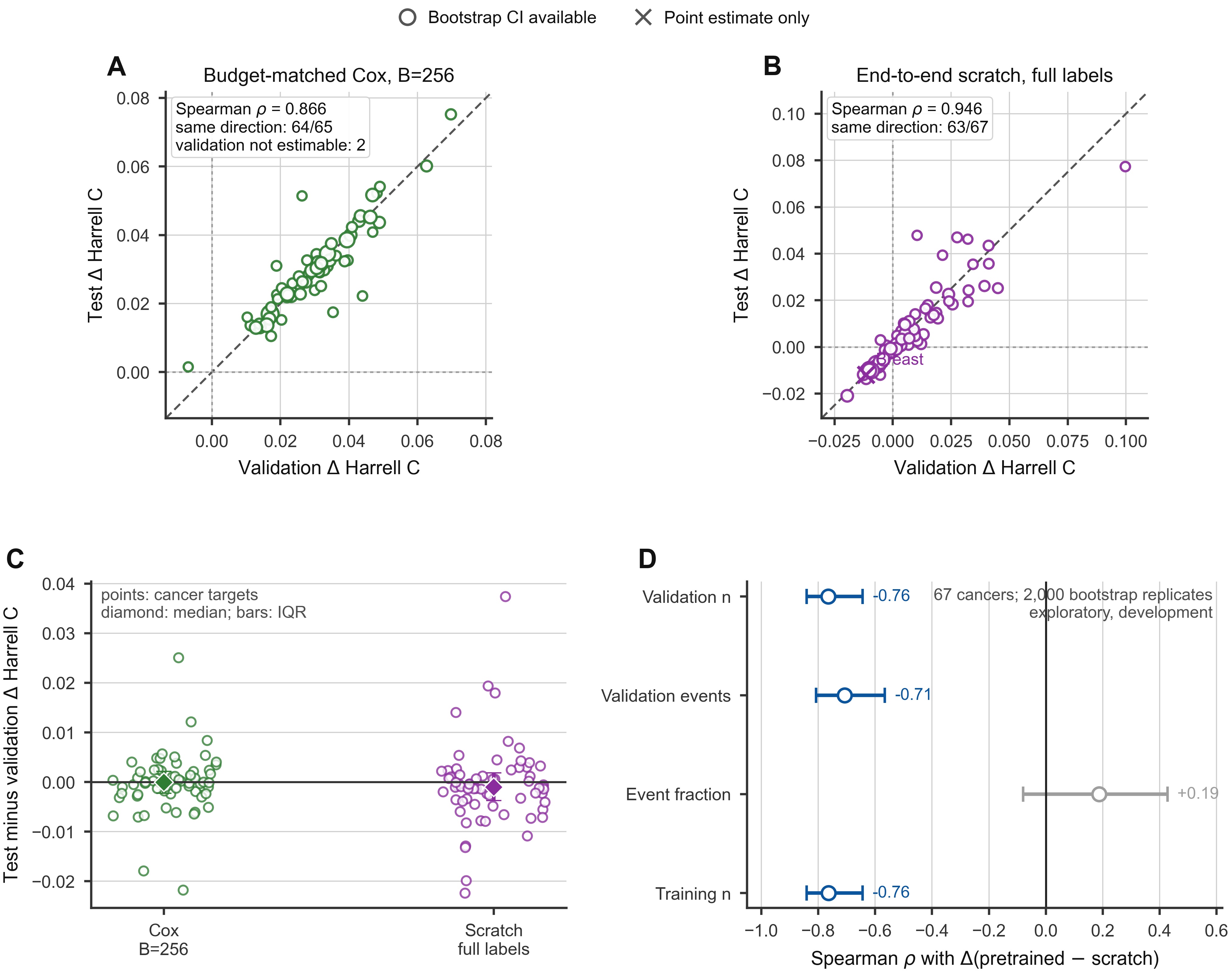

### Supplementary Figure S5

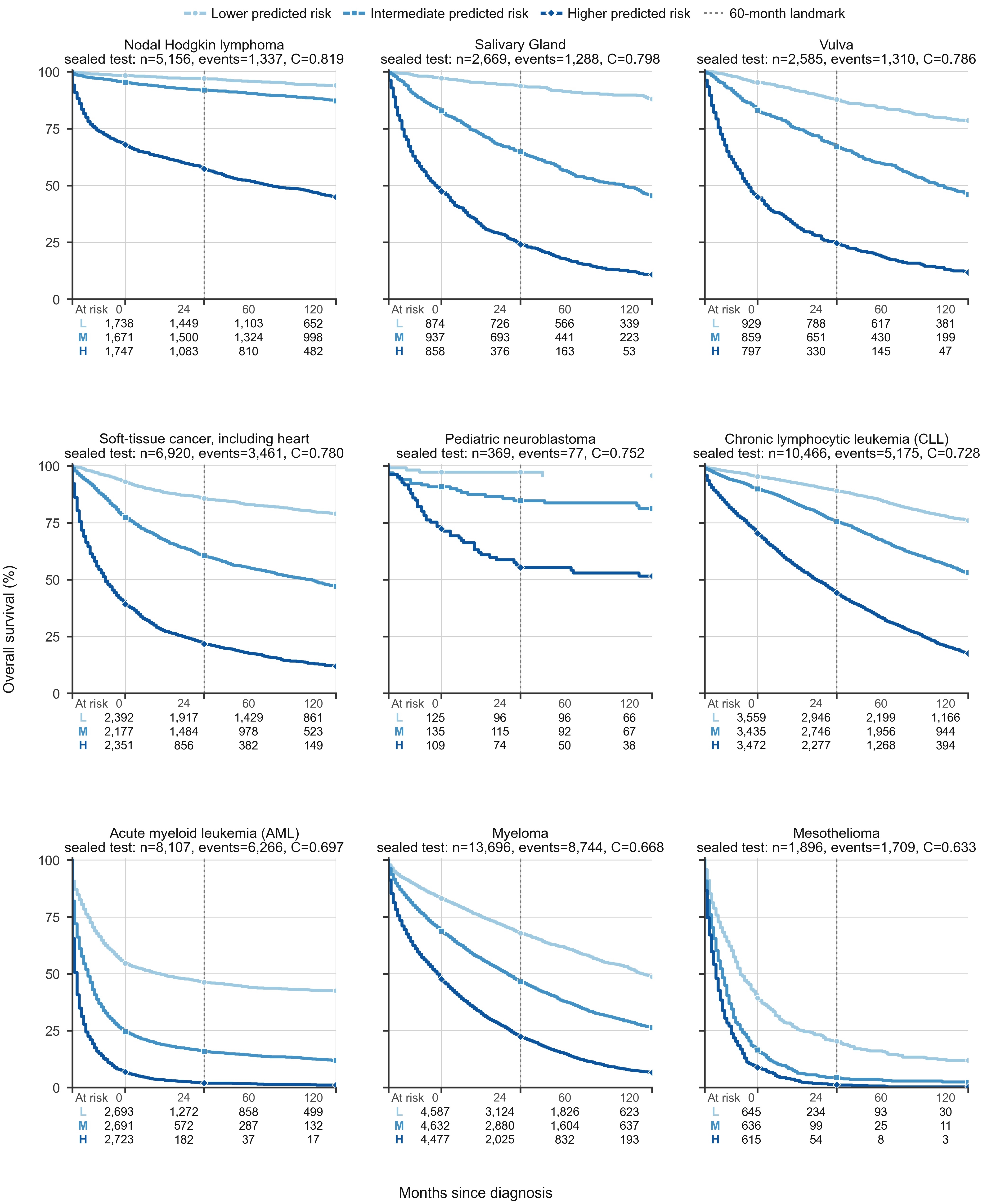

### Supplementary Figure S6

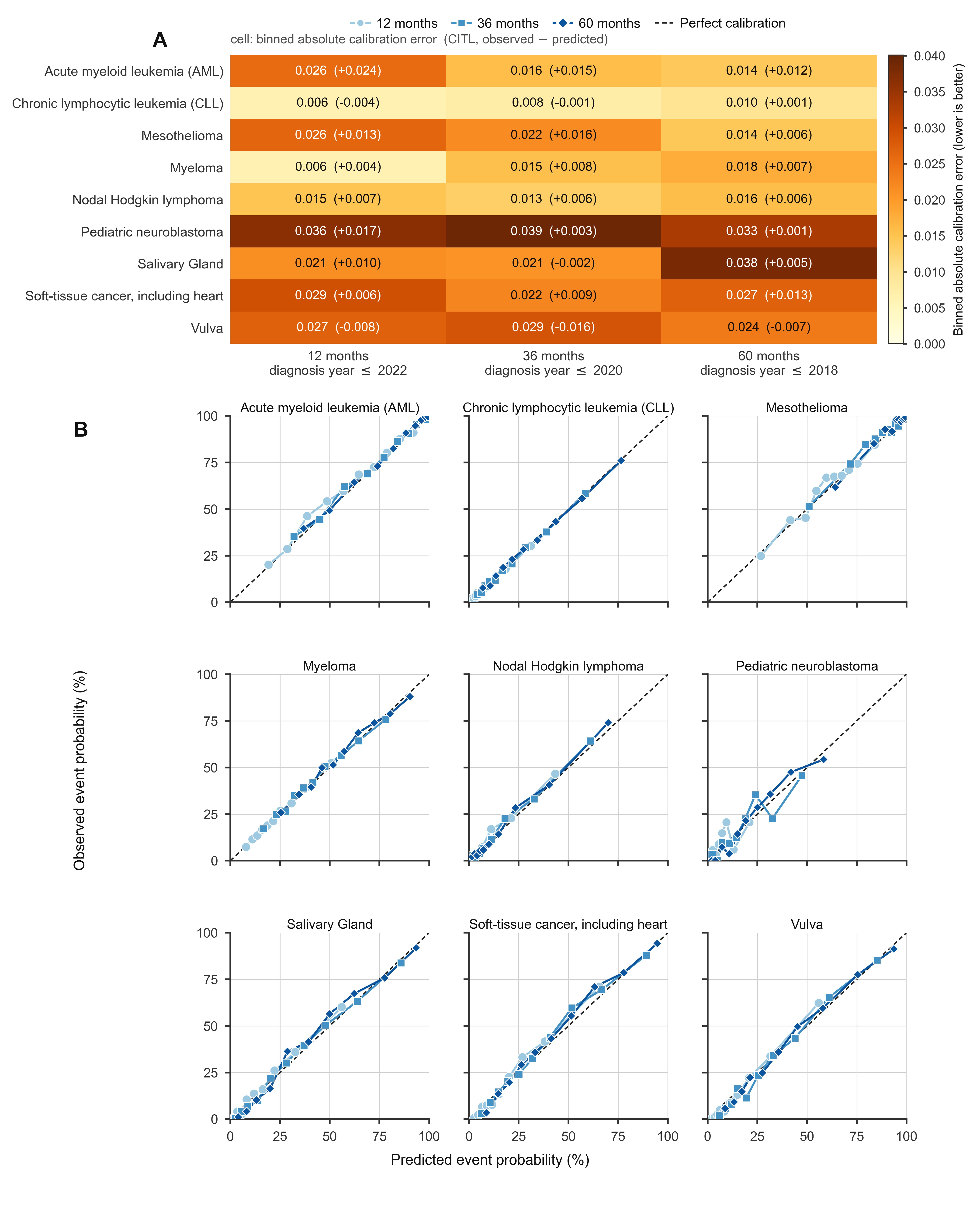

### Supplementary Figure S7

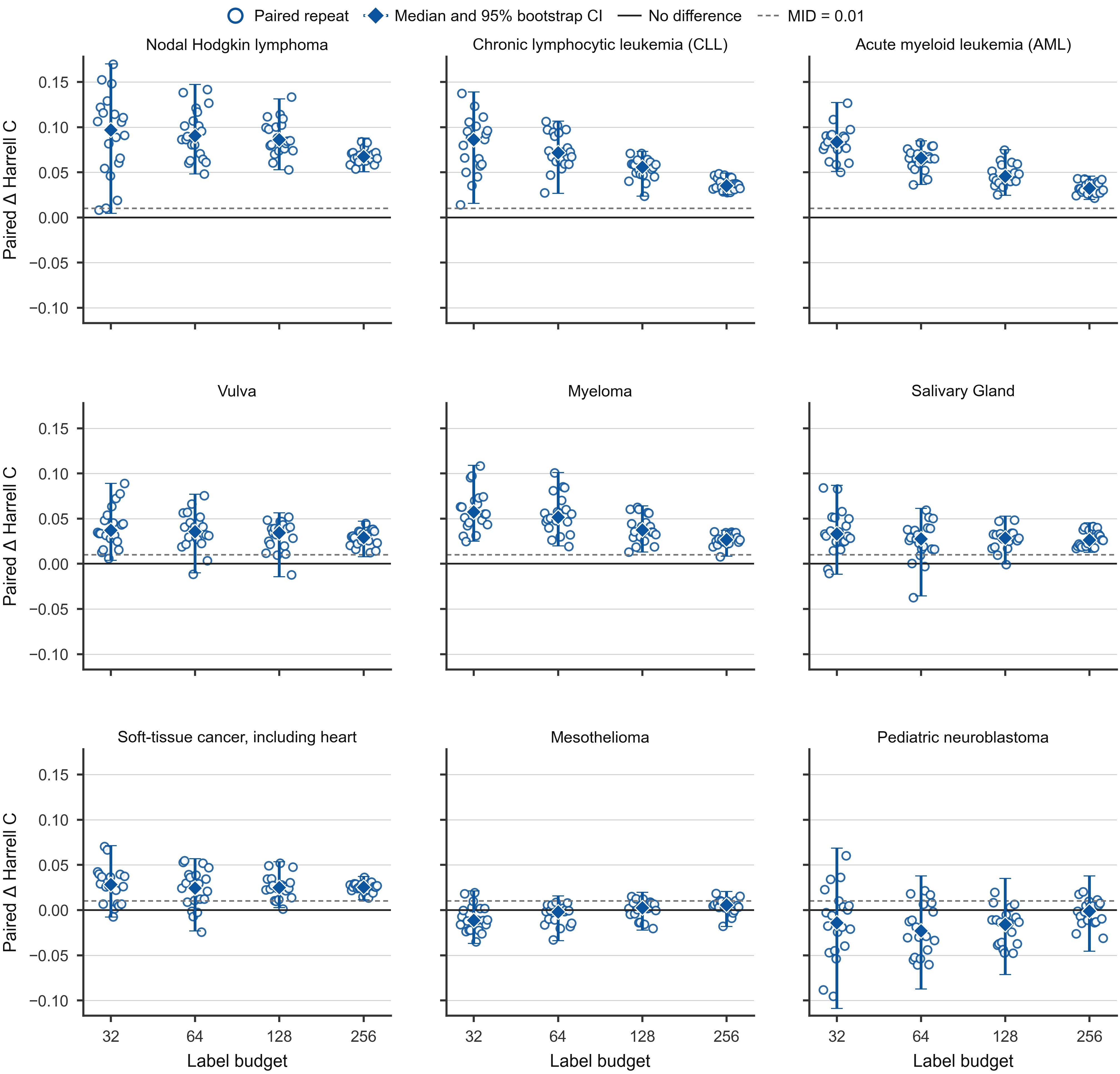

### Supplementary Figure S8

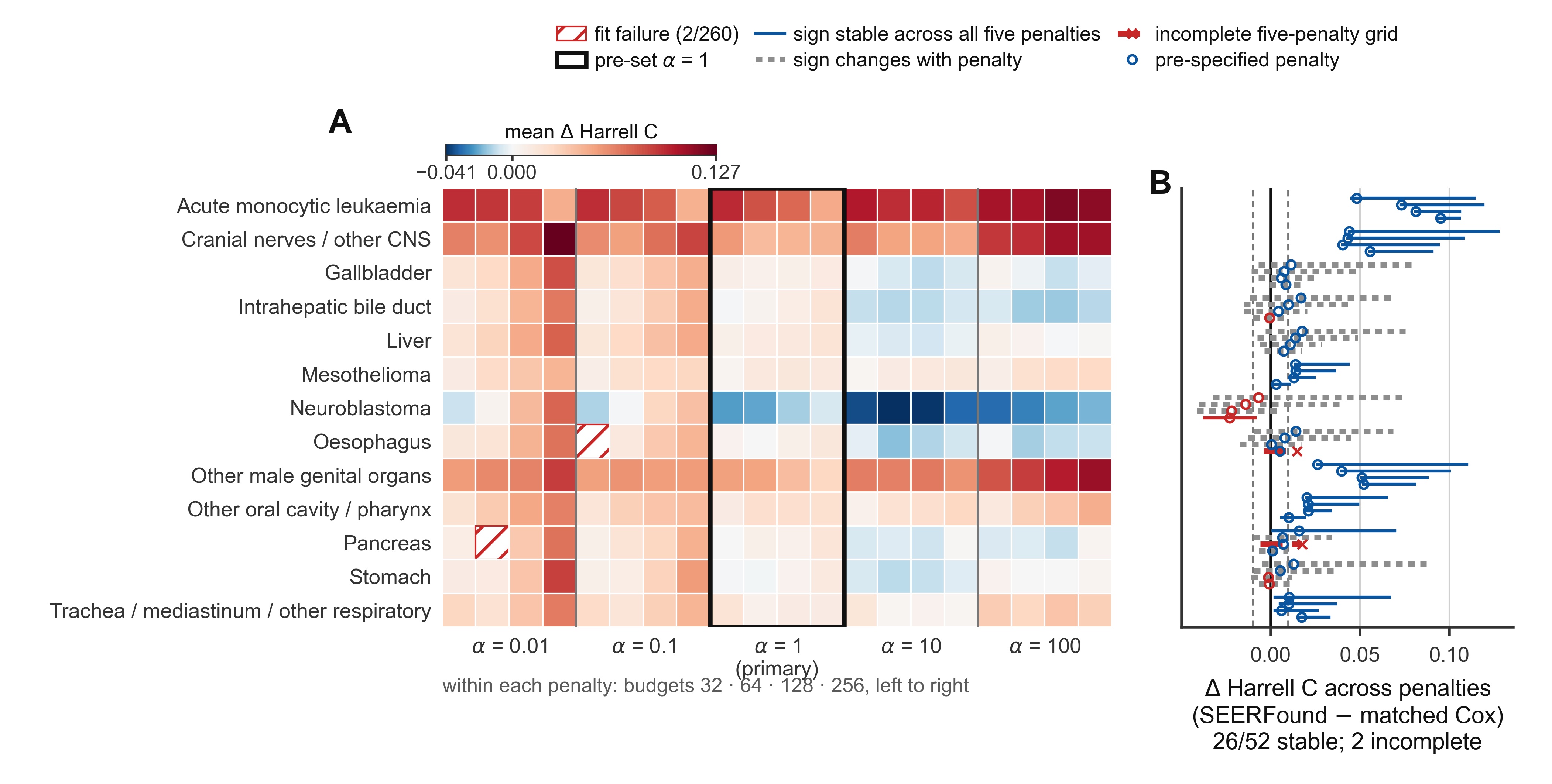
